# Functional Magnetic Resonance Imaging of the amygdala and subregions at 3 Tesla: A scoping review

**DOI:** 10.1101/2023.03.26.23287768

**Authors:** Sheryl L. Foster, Isabella A. Breukelaar, Kanchana Ekanayake, Sarah Lewis, Mayuresh S. Korgaonkar

## Abstract

The amygdalae are a pair of small brain structures, each of which is composed of three main subregions and whose function is implicated in neuropsychiatric conditions. Functional Magnetic Resonance Imaging (fMRI) has been utilised extensively in investigation of amygdala activation and functional connectivity with most clinical research sites now utilising 3 Tesla (3T) MR systems. However, accurate imaging and analysis remains challenging not just due to the small size of the amygdala, but also its location deep in the temporal lobe. Selection of imaging parameters can significantly impact data quality with implications for the accuracy of study results and validity of conclusions. Wide variation exists in acquisition protocols with spatial resolution of some protocols suboptimal for accurate assessment of the amygdala as a whole, and for measuring activation and functional connectivity of the three main subregions, each of which contains multiple nuclei with specialised roles. The primary objective of this scoping review is to provide a comprehensive overview of 3T fMRI protocols in use to image the activation and functional connectivity of the amygdala with particular reference to spatial resolution. The secondary objective is to provide context for a discussion culminating in recommendations for a standardised protocol for imaging activation of the amygdala and its subregions. As the advantages of big data and protocol harmonisation in imaging become more apparent so, too, do the disadvantages of data heterogeneity.

## Introduction

Functional Magnetic Resonance Imaging (fMRI) has become one of the most powerful tools in the investigation of functional organisation of the brain since its inception in the early 1990s [1]. Continuous technical developments in this non-invasive technique have led to its broad adoption for investigating neural activation and functional connectivity (FC) within the brain [2, 3], particularly by neuropsychiatric researchers investigating potential neural alterations in mental health conditions [4].

From a neuroimaging standpoint, FC has been defined by Friston et al., as “…the temporal correlations between spatially remote neurophysiological events” [5]. Put simply, different areas of the brain are considered to be part of the same functional network, demonstrated through a statistical relationship, if they are ‘active’ at the same time [6].

One region that has been identified in structural and fMRI studies as being broadly implicated in many mental health conditions is the amygdala [7]. The amygdalae, a key component of the brain’s emotion circuitry [8], comprises a pair of very small almond-like structures located in the temporal lobes [9]. fMRI using both task and resting-state techniques has facilitated investigation of the functional role of the amygdala and how it is connected to the rest of the brain in mental health conditions [7, 10]. This is important as it has been posited that changes in the organisation of functional brain connections are responsible for mental health conditions rather than their causes being attributed to local or regional structural abnormalities [11].

Despite being the subject of much investigation, the role of the amygdala and its functional connections with other brain regions remains poorly understood [12]. This is due, in part, to its diminutive size with average volumes reportedly around 1240mm^3^ (1.24cm^3^) [13]. A further barrier to greater understanding is that the amygdala comprises nine functionally different nuclei which are grouped into three discrete subregions, the laterobasal (LB), centromedial (CM) and superficial (SF) (Fig. 1). It has been previously shown that each subregion has a specialised role and displays differential connections to the rest of the brain [14-17].

**Figure 1:**
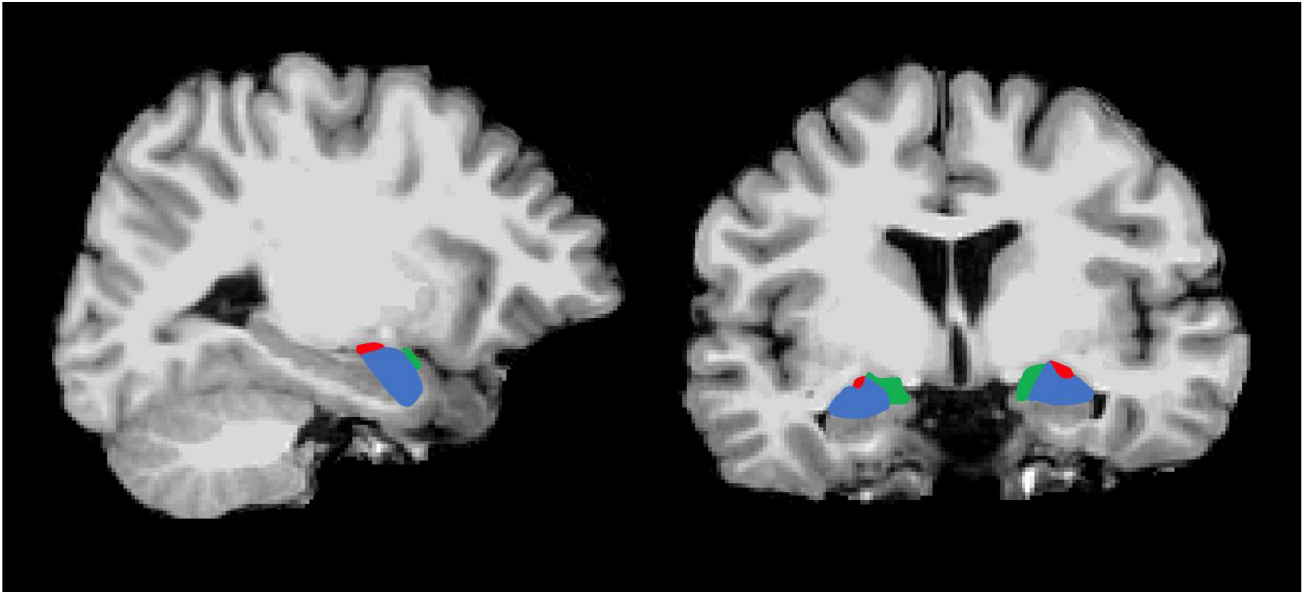
T1-weighted sagittal (left), and coronal (right) images showing location of the amygdala and 3 main subregions (SF = green, LB = blue, CM = red)

Blood Oxygenation Level Dependent (BOLD) contrast, based on an intrinsic sensitivity to local alterations in oxygen consumption driven by neural activation, is the primary mechanism used in fMRI to detect activation in the brain. [18]. BOLD signal changes detected on 1.5 Tesla (1.5T) and 3 Tesla (3T) systems are very small, being only 1-5% [19, 20] and Contrast-to-Noise ratio (CNR) provides a measure of these relatively small BOLD signal fluctuations compared to the noise. Aside from the modest BOLD signal changes achieved, a limitation to its use in investigation of the amygdala is the conflicting requirement for high resolution in both space (spatial resolution) and time (temporal resolution) [21]. Signal-to-noise ratio (SNR) is the ‘currency’ of MRI and is a major determinant of the levels of spatial and temporal resolution achievable. Typically, imaging sequences provide high spatial or high temporal resolution with each outcome being compromised by the other. As SNR and field strength are directly proportional, ultra-high field strength systems such as 7 Tesla (7T) and above possess inherently high SNR and can produce images with higher spatial resolution and larger activation-related signal changes [22, 23]. However, the bulk of fMRI clinical research is currently performed on 3T systems due to their relatively wider availability and, although the SNR levels are proportionally lower than 7T, there are fewer issues with problematic susceptibility and dephasing artifacts which are a function of ultra-high field strength systems [22]. 3T is now considered as the standard field strength for fMRI studies [24]; however, acquisition protocols appear to be variable, particularly in relation to spatial resolution, resulting in data that is potentially suboptimal in the investigation of very small structures [25-27].

High spatial resolution, indicated by voxel volume (VV) in mm^3^, is required in order to accurately image very small structures such as the amygdala and its subregions [28] and this level of resolution is easily achievable in structural MRI with VV of less than 1mm^3^ (0.001cm^3^) currently in common use [29]. However, for fMRI protocols, data is acquired at much lower spatial resolution, with larger VVs of 20-50mm^3^ considered as ‘standard’ [24] and up to 100 mm^3^ commonly in use [30]. This is due to additional temporal constraints on the acquisition of task and resting state fMRI data such as task design, adequate sampling of the brain’s haemodynamic response and sufficient SNR and CNR for desired brain coverage. Slices covering the whole brain (one volume) are acquired within one repetition time (TR) period of typically 2-3 seconds which is long enough to account for the brain’s relatively slow haemodynamic response [21] and multiple volumes form a time series dataset which can be used to estimate the haemodynamic response function for each task condition. Due to the direct proportionality between SNR and VV the combination of temporal requirements results in parameter selection compromises, making it more challenging to achieve higher spatial resolution [30]. Published fMRI studies reveal wide data heterogeneity with voxel volumes in use for reporting on amygdala activation and connectivity ranging from 8mm^3^ (0.008cm^3^) to 64mm^3^ (0.064cm^3^) [26, 27].

Given reports of additional activation and FC patterns being differentiated in many brain regions using high spatial resolution fMRI at both 3T and 7T [22, 23], optimisation and wider implementation of a high resolution technique at 3T is overdue, particularly for the amygdala and its significantly smaller subregions. Other aspects of the data collection process also deserve scrutiny. Appropriate selection of acquisition plane can potentially reduce through-plane signal dephasing and improve SNR levels [31, 32]. Similarly, the choice of radiofrequency (RF) coil design is an important consideration which can impact significantly on SNR levels and achievable spatial resolution [33].

Many technical choices are made in the establishment of fMRI protocols, with sites having differing access to hardware and software options and varying levels of expertise. However, selection of optimal VV is an important aspect that can be easily implemented with positive consequences for data quality [34]. The focus of this scoping review is on the spatial resolution values achieved in a wide range of published fMRI protocols used for reporting amygdala activation and FC. Its objective is to provide a comprehensive overview for the purpose of stimulating discussion in three key areas:

i. variations in VV used in studies evaluating activation and FC in the amygdala and its subregions
ii. potential advantages of protocol optimisation such as improvements in data quality and reproducibility with an emphasis on the amygdala subregions
iii. value of protocol harmonisation in addressing the challenges of data heterogeneity in big data collaborations

## Methods

This review protocol follows the Preferred Reporting Items for Systematic reviews and Meta-Analyses extension for Scoping Reviews (PRISMA-ScR) methodology [35] as well as the 5-step methodological framework of Arksey and O’Malley [36]. The scoping review protocol is registered at the Open Science Framework (osf.io/e3c28) and published as a preprint online [37].

Adhering to the recommendations of Bramer and colleagues [38] for achieving maximum recall rate, the following databases were searched on February 9^th^ and 11^th^, 2022; Medline, Embase, Web of Science, Google Scholar and Scopus. Medical Subject Headings terms for the search, which excluded non-human studies, encompassed the following terms:

○ Functional Magnetic Resonance Imaging OR functional MRI OR fMRI
○ Amygdal* OR amygdal* nucleus
○ Functional connect* OR FC (functional connectivity)
○ 3 Tesla OR 3T

The database searches yielded a total of 572 publications which were then imported into Covidence software [39]. Of these, 186 were automatically identified as duplicates leaving 386 for screening and data extraction. Two authors (SF and IB) independently performed all screening; 75 records were excluded during abstract and title screening whilst a further 119 were excluded at full-text review resulting in a total of 192 articles for inclusion. The mean number of participants per study was 69. A PRISMA flowchart outlining the search, screening and selection strategy along with reasons for exclusion is shown at Figure 2.

**Figure 2:**
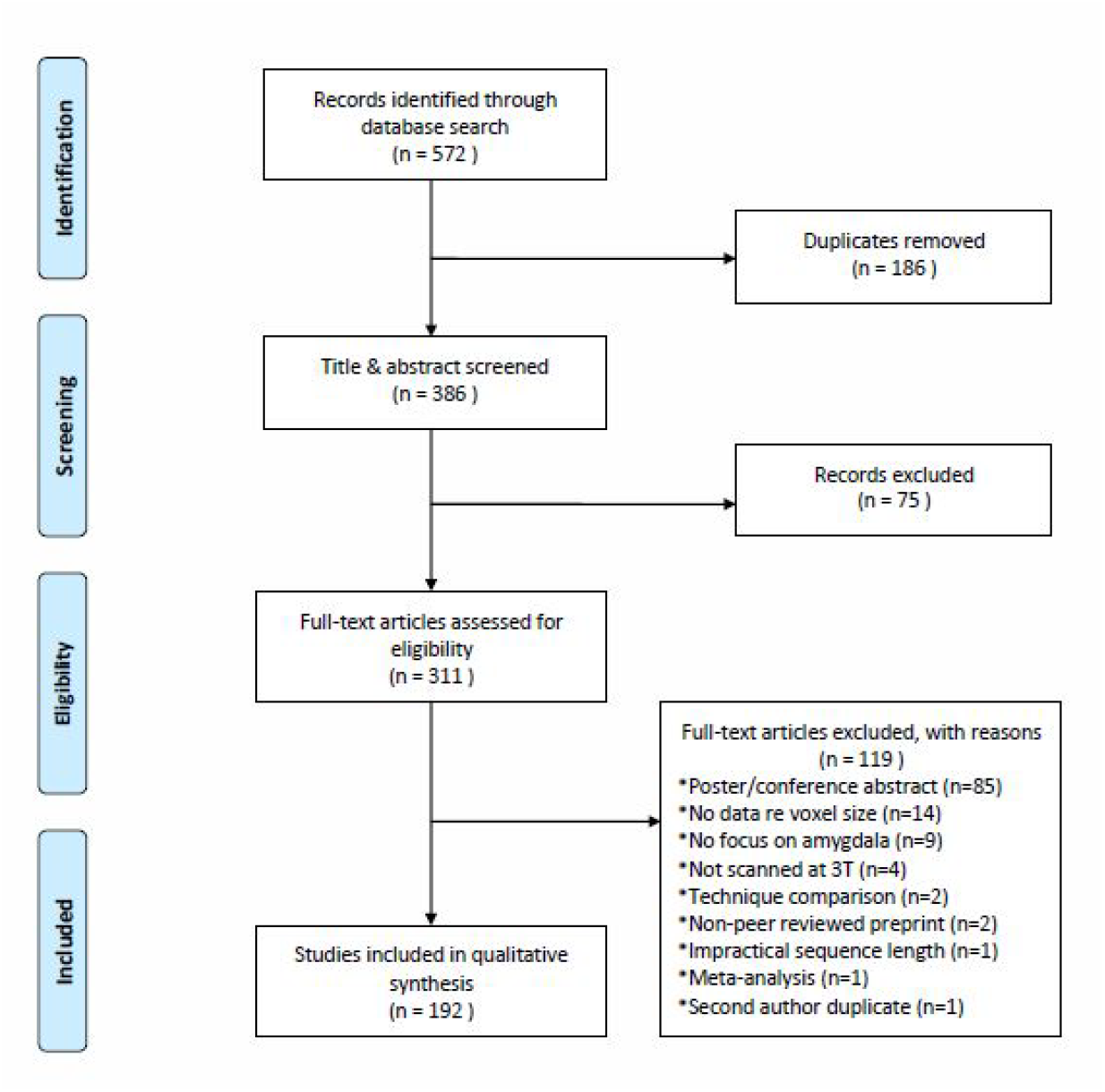
PRISMA flowchart outlining inclusion and exclusion process

Given that fMRI data quality and inherent image resolution is a complex topic, six sub-questions were investigated and data were extracted for discussion:

i. What sequence type was used for data acquisition?
ii. What spatial resolution values were achieved (VV in mm^3^)
iii. What imaging plane was utilised?
iv. Was whole brain coverage achieved?
v. What was the sequence acquisition time?
vi. What type of radiofrequency (RF) coil was utilised for signal reception?

## Overall Results

As the scope of this review captured a large number of publications the full data extraction table and reference list is provided in the Supplementary materials.

Table 1 at page 25 provides a summary of all extracted data (n=192).

**Table 1:** Data summary of all studies (n=192)

| Sequence Type |  | Spatial Resolution |  | Imaging plane |  | Full brain coverage |  | Acquisition time (TA) |  | RF Coil Type |  |
| --- | --- | --- | --- | --- | --- | --- | --- | --- | --- | --- | --- |
|  |  | VV range = 7.3mm<sup>3</sup> to 100mm<sup>3</sup>;<br>Slice thickness (ST) range = 2mm to 5mm |  |  |  |  |  | TA range = 2m 44s to 24 m |  |  |  |
| GE-EPI | 175 | High Res 1-20mm<sup>3</sup> | 30 | Axial/axial oblique | 186 | Yes | 185 | < 6 min | 23 | 8 Channel | 35 |
| Spiral version | 8 | Std Res > 20-50mm<sup>3</sup> | 126 | Coronal/coronal oblique | 2 | No | 7 | 6 to 8 min | 52 | 12 Channel | 21 |
| GE-EPI & Spiral | 3 | Low Res > 50mm<sup>3</sup> | 29 | Sagittal | 4 |  |  | > 8 to 10 min | 21 | 16 Channel | 1 |
| SE-EPI | 2 | Mixed Resolution | 7 |  |  |  |  | > 10 min | 13 | 20 Channel | 1 |
| DE-EPI | 1 |  |  |  |  |  |  | Mixed TA | 4 | 24 Channel | 1 |
| Gradient SE | 1 | ST ≤3.5mm | 127 |  |  |  |  | Not Stated | 79 | 32 Channel | 42 |
| ASL | 1 | ST >3.5mm | 65 |  |  |  |  |  |  | 64 Channel | 3 |
| GE-EPI & ASL | 1 | No Slice Gap | 143 |  |  |  |  |  |  | Mixed | 3 |
|  |  | Slice Gap | 45 |  |  |  |  |  |  | Other | 17 |
|  |  | Mixed gap/no gap | 4 |  |  |  |  |  |  | Not Stated | 68 |

### i. Sequence type

The T2*-weighted two-dimensional (2D) Gradient Echo-Echo Planar Imaging (GE-EPI) sequence was the most common acquisition sequence with 85% of studies (n=164) reporting its use exclusively and a further 6% (n=11) of authors not specifically naming their sequence but reporting parameter selections in keeping with GE-EPI. A total of 11 studies used spiral sequences, three of which also used GE-EPI and combined the data from both sequences. Two studies used Spin Echo-Echo Planar Imaging (SE-EPI), one used dual-echo EPI, one used a gradient spin echo technique and one used the Arterial Spin Labelling (ASL) technique. One study acquired data with both GE-EPI and ASL techniques.

A majority of studies (56%, n=108) focused on acquisition of resting state fMRI (rs-fMRI) data while 35% (n=68) focused on task-based fMRI, where participants are asked to view images or actively engage in a task during the scan. Sixteen studies reported both task and rs-fMRI acquisitions, whilst two studies by the same group reported using task data for intrinsic resting state data analysis [40, 41], an approach that has been previously validated [42].

### ii. Spatial resolution

VV ranged widely from 7.3mm^3^ (pixel size =2.44mm^2^ x slice thickness=3mm) to 100mm^3^ (pixel size = 25mm^2^ x slice thickness = 4mm). Fourteen of the 192 studies aggregated data from multiple protocols and scanners, only one of which used the same VV across protocols. To facilitate presentation and discussion of our results, we used Olman and Yacoub’s proposal of VV ranges. From 1mm^3^ to 20mm^3^ was considered high resolution and greater than 20mm to 50mm^3^ was considered standard resolution [24]. All studies with VV greater than 50mm^3^ were considered to be low resolution.

Of the 192 studies reviewed, only 16% (n=30) acquired high resolution data exclusively with a further four combining high and standard resolution data. All data acquired in 66% of studies (n=126) was standard resolution with a further three combining standard and low resolution data. Of the remaining studies, 15% (n=30) reported use of low resolution data exclusively and a further three combined low and standard resolution data. Of the low resolution studies, the data from all but one fell into the range of 50-70.3mm^3^ with a single study reporting a VV of 100mm^3^.

Slice thickness or through-plane resolution ranged from 2mm to 5mm with 66% of studies (n=127) exclusively using data with the median slice thickness of 3.5mm or less. The protocol in 74% of studies (n=143) did not use a slice gap technique. Studies not specifically reporting a slice gap were presumed to have none. Of the remaining 49 studies reporting a slice gap, four combined data using both techniques.

Slice gaps, commonly referenced as a percentage of slice thickness, ranged from 0% - 33.4% of slice thickness in 88% of the 49 studies using this technique. Of the remainder, two studies employed a gap of 50% (4mm/2mm, 2mm/1mm), two used a gap of 53.8% (2.6mm/1.4mm), one used a gap of 100% (3mm/3mm) whilst one reported a gap of 125% (3.4mm/4.25mm).

### iii. Imaging Plane

Axial or axial oblique acquisition planes were by far the most popular choice, with 97% of studies (n=186) utilising this plane. Of the remaining six studies, four used sagittal and two used coronal or coronal oblique planes. Studies not directly reporting an acquisition plane (n=68) were presumed to have acquired axial or oblique axial slices due to this being the most commonly reported plane [43].

### iv. Whole brain coverage

Whole brain coverage was obtained in 96% of studies (n=185), with all but six of these also utilising the axial or oblique axial plane. Of the seven studies acquiring limited brain coverage, only five acquired high resolution data and all used axial or oblique axial planes as well.

### v. Sequence acquisition times

There was a wide variation in acquisition times (TA) with the shortest sequence being reported at 2 minutes 44 seconds and the longest single TA at 24 minutes. TA was unreported in 41% of studies (n=79) and, of the remaining 113 studies, 20% (n=23) used TA of less than 6 minutes only, 46% (n=52) used a TA of 6 to 8 minutes only, 19% (n=21) used a TA of more than 8 to 10 minutes only and 12% (n=13) used a TA greater than 10 minutes only. Four studies reported TAs from two different TA groups, due to either the use of different TA for task and rs-fMRI acquisitions or the combination of data from different sites and or MRI systems. Two studies combined four separate 15 minute acquisitions for a total of one hour of data.

### vi. Radiofrequency Coils

As expected, most studies used data acquired on a single coil type with the most commonly used being the 32 channel phased array in 22% (n=42) of studies. This was followed by the 8 channel phased array with 18% (n=35) and 12 channel phased array with 11% (n=21). Three studies utilised the newer technology 64 channel head/neck phased array whilst one used the 16 channel phased array, and one each used 20 and 24 channel phased arrays. Three studies combined data acquired on different coil types, two of which used data from both 8 and 32 channel phased arrays whilst one combined data from 12 and 16 channel phased arrays. The remaining 9% (n=17) reported use of birdcage, quadrature, custom-designed or other ambiguously-named coil types. In approximately 35% of studies (n=68) the authors reported no information on RF coil type.

## Results for studies reporting subregional findings

Twenty one studies reported subregional activation in the amygdala [17, 44-63]. A summary of the data relating to these studies is reported in Table 2 (attached). Of these, eighteen used the GE-EPI sequence and one each used the spiral and DE-EPI techniques, with one combining data acquired with both GE-EPI and ASL techniques. Only four studies used high resolution data exclusively [17, 45, 46, 62] whilst two other studies used a combination of high and standard resolution data [59, 64]. Ten studies used standard resolution data [47-49, 53-56, 61, 63, 65]. Four studies used low resolution data [51, 52, 57, 60] and one combined standard and low resolution data [50]. Interestingly, four studies reported subregional findings using low resolution data with VV larger than 50mm^3^ [51, 57], two of which used VV of 70.3mm^3^ [52, 60]. Four of the studies used a slice gap technique, two of which used a gap of 33.3% [54, 55], one used a gap of 25% [50] and one used a gap of 12.5% [56].

**Table 2:** Data summary of subregional studies (n=21)

| Sequence Type |  | Spatial Resolution |  | Imaging plane |  | Full brain coverage |  | Acquisition time (TA) |  | RF Coil Type |  |
| --- | --- | --- | --- | --- | --- | --- | --- | --- | --- | --- | --- |
|  |  | VV range = 8-70.3mm<sup>3</sup><br>Slice thickness (ST) range = 2-5mm |  |  |  |  |  | TA range = 4m 48s to 15m |  |  |  |
| GE-EPI | 18 | High Res 1-20mm<sup>3</sup> | 4 | Axial/axial oblique | 20 | Yes | 21 | < 6 min | 2 | 8 Channel | 1 |
| Spiral version | 1 | Std Res > 20-50mm<sup>3</sup> | 10 | Axial oblique (-30 degrees) | 1 | No | 0 | 6 to 8 min | 8 | 12 Channel | 2 |
| DE-EPI | 1 | Low Res > 50mm<sup>3</sup> | 4 | Coronal/coronal oblique | 0 |  |  | > 8 to 10 min | 2 | 32 Channel | 7 |
| GE-EPI & ASL | 1 | Mixed Resolution | 3 | Sagittal | 0 |  |  | > 10 min | 4 | Mixed 12 & 16 channel | 1 |
|  |  | ST ≤ 3.5mm | 11 |  |  |  |  | Not Stated | 5 | Other | 3 |
|  |  | ST > 3.5mm | 8 |  |  |  |  |  |  | Not Stated | 7 |
|  |  | Mixed slice thickness | 2 |  |  |  |  |  |  |  |  |
|  |  | No Slice Gap | 17 |  |  |  |  |  |  |  |  |
|  |  | Slice Gap | 2 |  |  |  |  |  |  |  |  |
|  |  | Mixed gap/no gap | 2 |  |  |  |  |  |  |  |  |

All 21 studies acquired whole brain data in the axial oblique plane with one study reporting use of a ‘-30 degree’ slice tilt to minimise signal loss [63]. TA ranged from 4 minutes 48 seconds in a study using VV of 42.9mm^3^ [47] to 15 minutes per sequence in two studies that utilised Human Connectome Project [66] data with VV of 8mm^3^. These groups combined data from four 15-minute acquisitions for analysis [17, 45]. Five studies did not report their TA but of the 16 that did, 50% used a TA from 6-8 minutes. Seven studies did not report on their RF coil type, but of the remaining 14, 50% used a 32-channel phased array.

Table 2 at page 26 provides a data summary of subregional studies (n=21)

## Discussion

In providing an overview of the acquisition protocols in use at a large number of imaging sites, this comprehensive review has revealed the disparity in spatial resolution of fMRI data acquired to investigate the amygdala and its subregions. Whilst the same sequence type and imaging plane was used in a large majority of studies, spatial resolution values and acquisition times were found to vary widely. This was particularly notable in the studies reporting subregional activation in the amygdala, with results in 81% of these studies relying on standard, low or mixed resolution data. This use of suboptimal techniques to image the amygdala together with significant variation in scanning parameters may be contributing to incongruent findings across studies as well as lack of success in study replication.

### Sequence of choice for BOLD signal at 3T

The T2*-weighted Gradient Recalled Echo (GRE) or Gradient Echo (GE) pulse sequence is inherently sensitive to BOLD-related susceptibility effects and is the mainstay in use at 3T due to the predominance of T2* contrast over T2 contrast [3]. The 2D version of the GE Echo Planar Imaging (GE-EPI) sequence, with its high BOLD sensitivity, is considered the gold standard [43]. In addition, its ease of implementation and wide availability in both research and clinical environments has rendered it the sequence of choice for fMRI of the brain [24] and this was evident in our review with over 90% of studies employing this technique. The primary difference between spiral and GE-EPI sequences is the k-space trajectories; spiral sequences traverse k-space from the centre to the periphery whilst GE-EPI employs a Cartesian trajectory of line-by-line acquisition [67]. Spiral k-space trajectory techniques have been shown to be superior to GE-EPI techniques in recovering signal in areas typically affected by susceptibility-induced field gradients such as frontal and medio-temporal areas, with the difference in magnetic susceptibility values at air/tissue interfaces typically being around 8 parts per million (ppm) [68]. However, the use of these techniques is not widespread, as evidenced in our review, partly due to the requirement for lengthy non-standard offline image reconstruction which many clinical research sites cannot support [69]. The use of alternative acquisition techniques was also minimal due to their shortcomings. Arterial spin labelling is limited in fMRI due to its relatively poor functional contrast and temporal resolution, despite its high spatial accuracy [70]. Gradient spin echo techniques are also not commonly used due to their lower sensitivity, reduced brain coverage and greater heat deposition into the patient reported as Specific Absorption Rate (SAR) in watts per kilogram [43].

### Data quality – SNR, CNR and spatial resolution considerations

Both SNR and CNR are used as measures in assessment of overall data quality in fMRI. As data provides information about signal fluctuations and stability over a time course SNR is referenced as temporal SNR (tSNR), or mean signal intensity over time, [25, 71]. As previously outlined, SNR and VV are directly proportional [19]. In theory, the use of smaller VV to achieve higher resolution fMRI data results in reduced tSNR per individual voxel which seems intuitively undesirable. Therefore in the compromise between tSNR and achievable spatial resolution when selecting VV, often spatial resolution suffers.

However, there are several other issues to consider in relation to overall data quality when choosing VV. Of primary concern is the location of the amygdala itself. tSNR is diminished by localised main magnetic field (B0) inhomogeneity which results in signal loss and image distortion at air/tissue interfaces. Intravoxel dephasing or signal drop-out occurs when B0 is inhomogeneous within an individual voxel, effectively reducing its SNR contribution [72]. Areas particularly impacted are the inferior temporal regions and sphenoid sinuses adjacent to the amygdala where protons are effectively spinning at different frequencies [73]. Also, with Cartesian k-space trajectories such as that used in GE-EPI sequences, distortion results from voxels being mismapped due to phase errors. In these scenarios, increasing spatial resolution by reducing VV has been shown to improve data quality [21] and potential increases in BOLD signal can be obtained by acquiring smaller voxels in areas such as air/bone interfaces that are traditionally difficult to shim well [19]. Similarly, Olman and colleagues reported that although tSNR was reduced when using smaller VV there was less signal variability across the Medial Temporal Lobe, resulting in more consistent BOLD sensitivity across the region of interest [72]. Further, Robinson et al., focusing their measurements on the amygdala, showed that the SNR penalty incurred in halving the VV by reducing the slice thickness to 2mm was offset by a notable decrease in intravoxel dephasing [74].

Partial volume averaging occurs with larger voxels that encompass a mix of tissue types and its effects can be reduced by matching the VV or spatial resolution to the size of the potential activation during the acquisition in order to maximise functional CNR [75]. Partial volume averaging effects have also been shown to be particularly significant in imaging the amygdala, not only due to its small size but its location adjacent to the Basal Vein of Rosenthal (BVR) which has been shown to give rise to confounding vascular signals when it is partially contained when using larger voxels [32].

As BOLD signal changes are effectively the signal of interest, CNR has been suggested as the primary indicator of data quality [20]. CNR is maximised by reducing noise contributions which have a greater negative impact in fMRI. Unlike background noise in MRI images, noise in fMRI time series data is a blend of physiological noise such as respiratory and cardiac pulsation and thermal noise being emitted by the MR system components as well as from the patient’s body [20, 76]. Maximal CNR has been shown to be achievable with higher spatial resolution imaging in conjunction with mitigation of noise contributed by artifactual and physiological signal fluctuations [19]. Physiological noise contributions also reduce tSNR levels and can be addressed during data post-processing and analysis [77].

Taken together, these technical points provide a solid basis for utilising smaller VV in fMRI focusing on the amygdala and it was somewhat surprising to find that only 16% of studies in this review had utilised high resolution data in reporting their findings.

### Selection of slice orientation and acquisition method

Data acquisition can be performed sequentially or in an interleaved fashion. Researchers typically use an interleaved slice acquisition technique which is advantageous as is allows the use of a shorter repetition time (TR) and precludes the use of a slice gap [78]. Using a sequential slice acquisition method without a slice gap can result in slice excitation leakage or adjacent slice saturation leading to reduced tSNR [79]; however, MR system manufacturers have developed and implemented methods to mitigate these effects [80]. Of the studies reporting use of a slice gap technique, it is unknown whether an interleaved or sequential slice acquisition technique was used as this data was not extracted for the current review.

A general disadvantage of using a gap technique is that spin-history artifacts can occur with through-plane head motion which is more severe with increasing gap size. On the other hand, the gap technique can be advantageous as it allows increased brain coverage whilst keeping within a defined TR and slice envelope. However, when imaging the amygdala, use of a gap technique could result in missing potential activation occurring in tissue in between the imaged slices [81-83]. As the almond-shaped amygdala is very small at only around 10mm in width [32] with an average volume of approximately 1240mm^3^ [13], the acquisition of interleaved contiguous axial 3mm slices would likely result in amygdala tissue appearing in only 3-4 slices, and even fewer if thicker slices or a slice gap technique was used. This highlights the importance of considering the size of the structure of interest when making protocol decisions about spatial resolution and slice acquisition techniques. Of the studies in this review reporting on the subregions 20% used some or all data acquired with slice gaps, two of which used standard resolution data with 3mm slices and a 33% gap whilst another used combined standard and low resolution data with 5mm slices and a 25% gap.

Slice orientation has the potential to contribute to signal loss in T2*-weighted imaging and selection of acquisition plane should be also considered in relation to the particular brain regions under investigation. The plane introduced by Talairach and commonly referred to as the AC-PC (Anterior Commissure – Posterior Commissure) line is reportedly the most widely used and has been broadly adopted as a standard reference angulation in fMRI acquisitions [43, 84] and this was evident in our review with 97% of studies reporting its use. Acquisition of whole brain data parallel to the AC-PC line is more time-efficient as it requires around 30% less slices compared to the coronal plane [74, 84]. The use of axial slices angled at approximately -30 degrees (that is, with the anterior edge of the slice tilted cranially) has been reported to improve BOLD sensitivity in the region of the amygdala as well as decreasing susceptibility-related signal loss; however, this angulation may be suboptimal for imaging other regions when acquiring whole brain data that is a requirement for FC studies [63, 85]. Only one study in our review utilised this slice angulation and they reported subregional findings using standard resolution data. Although the study was based on task activation rather than investigation of FC, whole brain coverage was acquired with this angulation [63]. Selection of imaging plane in relation to orientation of the structure of interest, especially when using rectangular or anisotropic voxels, is also an important consideration. The use of large voxels that are also anisotropic has been shown to provide inaccurate data on volume and shape of small nuclei as well as a reduction in signal sampling accuracy which is based on the largest voxel dimension. Therefore small isotropic voxels are preferred because of their cubic nature [86, 87].

Earlier studies on 1.5T systems have explored the use of other slice orientations for improved imaging of the amygdala. Merboldt et al., found that optimal BOLD imaging for investigation of the amygdala required data acquisition in the coronal plane [88]. Chen and colleagues also reported that, for the amygdala, the largest susceptibility field gradients occur in the superior-inferior (SI) direction and higher BOLD sensitivity was demonstrated in oblique coronal images in conjunction with frequency-encoding in the SI direction [89]. This was in agreement with Bandettini who noted the use of coronal and sagittal slice acquisitions in many studies for the same reason [19]. However, along with the arrival in the mid-2000s of 3T systems with more powerful gradients, came a number of additional technical considerations. Whilst superior gradient performance has many advantages, use of stronger gradients was restricted when using a coronal orientation in conjunction with SI phase encoding. This was due to reports of peripheral nerve stimulation and potential signal aliasing using this configuration, thereby limiting its use [34, 89]. A further consideration following the primary selection of phase encoding direction is the secondary choice, for example, SI or inferior-superior (IS). In areas affected by susceptibility artifacts this choice has been shown to impact on the accuracy of data analysis. Depending on the direction selected, images can be either distorted by distraction or compression; choosing the direction resulting in signal distraction is preferred as signal within regions of interest that have been compressed cannot be recovered during analysis [72]. In our review only four studies used the sagittal plane and two used coronal or coronal oblique planes. None of the six reported subregional findings and all acquired whole brain coverage with standard or low resolution VV, a protocol decision potentially driven by the conflicting need to acquire more slices for high resolution data when scanning in these orientations.

### Whole brain coverage versus a targeted approach

To accommodate the need for high spatial resolution data acquisition, multi-resolution approaches have been proposed in which a lower-resolution whole brain dataset is acquired to identify the site of activation, following which a targeted high-resolution dataset is acquired at that site [75]. In cases where the location of the target is known, it can be time-saving to focus only on the desired area of activation rather than acquiring high resolution data over the whole brain. The latter approach necessitates an increased number of slices for coverage resulting in increased TR and reduced temporal resolution. These issues present a conundrum when considering the optimal acquisition strategy for the amygdala as both high spatial and temporal resolution are required. If the purpose of the study is to investigate FC, whole brain coverage is necessary as activation needs to be interpreted within the context of amygdala connections with other spatially remote brain regions, rendering a reduced-coverage approach impractical [34, 90]. Seven studies in our review used a targeted approach with all acquiring task rather than resting-state data. Of these, five acquired high resolution data and two acquired standard resolution data. Only one of the 21 studies reporting on the amygdala subregions used a targeted approach and this group acquired high resolution data [62].

### Optimal sequence acquisition time

Choice of TA can involve compromise for a variety of reasons, with participant population being a major consideration. TA can also differ between task and resting state acquisitions with longer TAs of 15-25 minutes being recommended for FC studies with greater reproducibility [91]. However, Van Dijk and colleagues showed that fMRI with TA of around six minutes was satisfactory for demonstrating FC patterns in the brain and that performing more runs reflected only marginally on data reliability [92]. Of the 113 studies in our review that reported their TA, only 15 used a TA of less than six minutes to acquire FC data and 40 studies used a TA of six to eight minutes. Acquiring more data over a longer TA allows the possibility of discarding portions of motion-affected data whilst still retaining enough temporal measurements for analysis. However, as extended scan times can result in patient movement, TAs of six to eight minutes provide a reasonable compromise.

In the absence of acceleration techniques (discussed below) TA is a relatively simple calculation of the product of TR, number of slices and number of volumes acquired. Temporal resolution in fMRI, effectively the time taken to acquire all slices once, (i.e. one TR) is negatively impacted when acquiring higher spatial resolution data as more slices are required for whole brain coverage. Maintaining the same TA could be achieved by a reduction in the number of volumes acquired [79]. However, the very small signal changes of only around 1-5% make BOLD imaging extremely challenging and decreasing both the number of volumes and the VV in order to maintain the TA would significantly reduce tSNR [25]. In fact, one method used to improve tSNR when seeking to image at higher resolution is to acquire a greater number of time points or volumes by scanning for longer, thus reaping the benefits of signal averaging; however, a doubling of scan time only increases SNR by a factor of √2 [93].

The critical nature of the relationship between TA and tSNR in detection of fMRI effect size has been demonstrated by Murphy et al., whose work outlined the value to researchers of prospectively determining the required effect size to a specific p-value. Using this information together with tSNR values they demonstrated that the TA appropriate to each fMRI experiment can be calculated and that an increase in tSNR can significantly reduce TA for a given effect size [25].

### Radiofrequency coil technology

The evolution of RF coil technology and in particular, the introduction of phased array coils has positively impacted SNR levels and data quality [93]. This review revealed use of a wide range of coil types, including traditional volume transmit/receive types through to a variety of multi-channel phased array receivers ranging from eight to 64 channels Multi-channel phased arrays have been shown to provide SNR benefits over traditional volume coils, in part due to their improved filling-factor which relates to coil fit with regard to the region of interest [93]. Of the studies in this review reporting their coil type, 84% used a multi-channel phased array for signal reception.

An early demonstration of the superiority of multi-channel coil technology in the anterior Middle Temporal Lobe was reported by Bellgowan et al. in a comparison between higher and standard resolution slices (2mm and 4mm) acquired with both an older style transmit/receive volume coil and a 16 channel array receiver. They reported significant improvements in tSNR and CNR when acquiring high resolution data but noted this was only possible with the use of the multi-channel array coil [94].

Since their introduction, multi-channel coils have been adapted to accommodate increasing numbers of individual elements and 64 channel phased array coils are now available. A less desirable function of coils with very high numbers of elements is a reduction in depth penetration in line with decreasing element size. Higher SNR is evident peripherally due to the close proximity of the arrays to the anatomy whilst signal drop-off may be seen centrally, resulting in signal non-uniformity across the FOV [93]. Correction algorithms to compensate for this are widely available on commercial MRI systems, the use of which is recommended in studies focusing on deeper sub-cortical structures such as the amygdala [95].

The most commonly used coil type in this review was a 32 channel phased array which is intuitively a good compromise between high SNR and adequate depth penetration for improved signal uniformity, an important consideration as the amygdala is a relatively deep brain structure. Interestingly, in a direct comparison between 32 and 64 channel phased arrays, a study by Keil et al., found that the 64 channel phased array outperformed the 32 channel phased array centrally as well as peripherally in SNR terms when acceleration factors of 3 or more were used. This was due to g-factor (noise amplification) improvements attributed to the reduced element size allowing better spatial distinction of the variation in signal intensities and this was also the case when the Simultaneous Multi Slice (SMS) technique was employed [96]. As the 64 channel head/neck array is relatively new technology, its use may become more widespread as sites look to upgrade their head coils in the future.

Phased array coils possess several advantages over earlier coil designs. Along with the capability for imaging larger fields-of-view with inherently higher SNR, they allow spatial localisation of signals. This provides an opportunity to reduce TA by employing acceleration techniques such as parallel imaging and SMS, a technique which effectively increases the sampling rate [93]. In general, differing coil sensitivity profiles in these arrays allow signals to be spatially localised, thus enabling data undersampling with a concomitant reduction in TA. Data can then be fully reconstructed based on individual coil sensitivity profiles. However, the main advantage of parallel imaging in fMRI is not image acceleration but improved data quality resulting from reduced signal dephasing due to shorter achievable echo train lengths, inter-echo spacing and optimal echo time (TE) selection. The primary disadvantage of its use is the accompanying SNR penalty as a result of undersampling [79, 97]. Even so, Schmidt and colleagues recommended its use in the Medial Temporal Lobes, prioritising the marked decrease in image distortion in this location over the potential loss of SNR and BOLD sensitivity [98].

Conversely, the primary advantage of the SMS technique is a reduction in temporal resolution. In this case, the variations in coil sensitivity profiles enable spatial differentiation of multiple slices which can be excited simultaneously. This allows the use of shorter TRs, meaning that either TA can be reduced or statistical significance improved by acquiring more data points [99]. Whilst the SMS technique does not attract a SNR penalty, its implementation may result in higher SAR levels [78].

## Further considerations

There are many other aspects relating to fMRI data acquisition and quality that are outside the scope of this review. Firstly, the choice of echo times (TE) is an important consideration. Shorter TEs may result in improved data quality due to a decrease in susceptibility-related signal loss due to dephasing. However, the penalty is reduced BOLD sensitivity which is optimised when the effective TE value approximates the T2* of the tissue being imaged (around 40-50 ms at 3T) [98]. T2* values also increase with improved spatial resolution [19]. Robinson and colleagues, in a comparison between high and standard resolution VV, showed that the measured T2* value in the amygdala doubled from 22ms to 43ms in the high resolution data set, approaching the cortical value of 52ms which was similar at both resolutions [74]. This has obvious implications for researchers using shorter TEs based on T2* values calculated from standard or lower resolution data acquired in the amygdala.

Secondly, the use of SMS allows significant reductions in TR, speeding up the acquisition but also resulting in lower SNR if the TR is lower than the T1 of the brain tissue being imaged. TR values of lower than 500ms reportedly result in a marked reduction in functional contrast levels [19]. This was confirmed by McDowell and Carmichael in their 3T investigation of event-related tasks in which they showed there was little advantage to using TR values below 800ms [100]. However, the potential benefits in rs-fMRI may outweigh the primary disadvantage of using typical TRs of 2-3 seconds used in task acquisitions, this being aliasing of respiratory and cardiac signal fluctuations into the lower frequency range associated with resting state fluctuations, confounding the data [99] [101]. However, when using short TRs that could result in tissue saturation, the flip angle should be optimised to the Ernst angle to maximise SNR, and this may also have the added benefit of lower SAR [78]. Further, it has been shown that consideration needs to be given to choice of multiband (MB) factor (the number of slices acquired simultaneously) when using SMS. To prevent saturation artifacts, a simple calculation must be made in which the total number of slices divided by the MB factor should result in an odd number [78].

Although data processing and analysis techniques are integral to this topic, their discussion is also outside the scope of this review. However, it should be noted that, as improvements are made in spatial resolution of acquisitions, consideration should be given to suitability of data pre-processing steps and analysis techniques in current use. For high spatial resolution datasets, tSNR and detectability of BOLD signal changes have been shown to be increased by spatial smoothing [102]. However, other aspects of the data processing and analysis pipeline may detrimentally affect the spatial resolution of the acquired data and researchers have called for an awareness to look beyond the use of conventional parameters when pre-processing high resolution data in order to avoid nullifying any benefits derived from its acquisition [25, 103].

## Recommendations

We conclude this review with hardware and protocol recommendations for fMRI of the amygdala and its subregions that may be easily implemented in clinical and research sites.

i. 3 Tesla MRI system and 32 or 64 channel phased array coil to accommodate parallel imaging and SMS techniques
ii. T2*-W 2D GE-EPI high resolution sequence with voxel volumes of 20mm^3^ or less (preferably 2-2.5mm isotropic)
iii. Contiguous interleaved axial oblique images parallel to the AC-PC line
iv. Whole brain coverage to facilitate FC analysis
v. TA of 6-8 minutes (or prior determination of effect size at given p-value to calculate required TA)
vi. MR system-specific non-uniformity correction algorithm
vii. TR value optimised for study objective (task vs resting state)
viii. TE value optimised for spatial resolution of acquired data

## Limitations

The field of fMRI is broad with many variations in data acquisition techniques in use. Our purpose was to map out an overview of current techniques with a focus on spatial resolution. The narrow confines of the review are a major limitation due to the interrelatedness of multiple aspects of the data acquisition and analysis process, outlined above, that fell outside its scope. However, it focuses on one non-trivial aspect of the acquisition process that can be easily manipulated by researchers looking to optimise their data to answer specific questions.

A further limitation is that articles in-press or published after our literature search are not included. However, our selection of databases and search strategy was designed to maximise the number of publications identified for inclusion.

Lastly, we have not attempted to compare SNR levels between studies as there are many complex factors determining these values for individual studies. Due to lack of consensus among researchers, both SNR and CNR values can be calculated using different methods resulting in different scales, rendering any comparisons meaningless [20].

## Conclusion

This scoping review provides an overview of the fMRI protocols in use for investigation of amygdala activation and connectivity. Data quality in fMRI is contingent on a complex amalgamation of components which includes both CNR and SNR and, by extension, spatial resolution. Whilst researchers are locked in to their site’s field strength and scanner capability and, to a lesser extent, RF coil and sequence availability, decisions relating to parameter selections such as voxel volume, acquisition time, slice orientation and coverage should be carefully scrutinised by researchers in the context of their study objectives. This review has identified considerable disparity in fMRI acquisition protocols across imaging sites, resulting in broad data heterogeneity, particularly in relation to studies focusing on the subregions. To counter this, we provide recommendations for optimisation of imaging the amygdala and its subregions at 3T. The continued focus on biomarker identification at an individual level in many health conditions, the advent of machine learning in large datasets and fewer funding opportunities for research has resulted in renewed interest in collaborative consortium-style approaches as demonstrated by groups such as ENIGMA (Enhanced Neuroimaging and Genetics through Meta-Analysis) and ICBM (International Consortium for Brain Mapping). Protocol optimisation and harmonisation across sites performing similar research studies would be a simple yet effective way to progress the field.

## Data Availability

All data produced in the present study are available upon reasonable request to the authors

## Data and code availability statement

The data are available from the corresponding author upon reasonable request.

## CRediT authorship contribution statement

**Sheryl L. Foster:** Conceptualisation, Methodology, Investigation, Writing - Original Draft, Writing - Review & Editing. **Isabella A. Breukelaar:** Methodology, Investigation, Writing - Review & Editing. **Kanchana Ekanayake:** Data Curation: Validation. **Sarah Lewis:** Conceptualisation, Writing - Review & Editing, Supervision. **Mayuresh S. Korgaonkar:** Conceptualisation, Writing - Review & Editing, Supervision.

## Acknowledgements

This work was supported by the Westmead Charitable Trust, Westmead Hospital. The authors would like to thank Elizabeth Haris for ctreation of T1-W images.

## Appendix. Supplementary materials

