## Supplementary material for "Functional Magnetic Resonance Imaging of the amygdala and subregions at 3 Tesla: A scoping review": Supplementary material_Table of publications and full data extraction.docx

**Supplementary material - Full list of all publications and data extracted**

| **First Author/Year** | **Title** | **Sequence type** | **Voxel volume (mm^3^)** | **Slice thickness/gap (mm)** | **Acquisition time (m:s)** | **RF head coil type** | **Imaging plane** | **Full brain coverage** | **FC of subregions reported** |
| --- | --- | --- | --- | --- | --- | --- | --- | --- | --- |
| [**Ahs *et al.* 2014**](https://pubmed.ncbi.nlm.nih.gov/23887817/) | Feature-based representations of emotional facial expressions in the human amygdala | Inverse spiral | 49 | 4/0 | 6m 30s | 8 channel | Axial/axial oblique | Yes | No |
| [**Aloi *et al.* 2021**](https://pubmed.ncbi.nlm.nih.gov/34730052/) | Altered amygdala-cortical connectivity in individuals with Cannabis use disorder | GE-EPI | 33.1 | 3.5/0 | 16m | 8 channel | Axial/axial oblique | Yes | No |
| [**Altinay *et al.* 2016**](https://pubmed.ncbi.nlm.nih.gov/27768670/) | Quetiapine Extended Release Open-Label Treatment Associated Changes in Amygdala Activation and Connectivity in Anxious Depression: An fMRI Study | GE-EPI | 21.9 | 3.5/0 | 5m 33s | 12-channel | Axial/axial oblique* | Yes | No |
| [**Ambrosi *et al.* 2017**](https://pubmed.ncbi.nlm.nih.gov/28369737/) | Insula and amygdala resting-state functional connectivity differentiate bipolar from unipolar depression | GE-EPI | 46.2 | 4/0 | 5m | no data | Axial/axial oblique* | Yes | Yes |
| [**Andreano *et al.* 2010**](https://pubmed.ncbi.nlm.nih.gov/20637290/) | Menstrual cycle modulation of medial temporal activity evoked by negative emotion | GE-EPI | 8 | 2/0 | 6m | no data | Axial/axial oblique | No | No |
| [**Andreescu *et al.* 2015**](https://pubmed.ncbi.nlm.nih.gov/24996397/) | Emotion reactivity and regulation in late-life generalized anxiety disorder: functional connectivity at baseline and post-treatment | GE-EPI for ASL | 100 | 4/2 | no data | no data | Axial/axial oblique | Yes | No |
| [**Anticevic *et al.* 2014**](https://pubmed.ncbi.nlm.nih.gov/24366718/) | Amygdala connectivity differs among chronic, early course, and individuals at risk for developing schizophrenia | GE-EPI | 42.2 | 3/0 | 6m 40s | 12-channel | Axial/axial oblique | Yes | No |
| [**Assaf *et al.* 2018**](https://pubmed.ncbi.nlm.nih.gov/29931835/) | Neural functional architecture and modulation during decision making under uncertainty in individuals with generalized anxiety disorder | GE-EPI | 46.2 | 3/1 | no data | no data | Axial/axial oblique | Yes | No |
| [**Avidan *et al.* 2014**](https://pubmed.ncbi.nlm.nih.gov/23377287/) | Selective dissociation between core and extended regions of the face processing network in congenital prosopagnosia | GE-EPI | 42.9; 19.1 | 3.5/0; 3.5/0 | 10m | Standard | Axial/axial oblique | Yes | No |
| [**Awasthi *et al.* 2020**](https://pubmed.ncbi.nlm.nih.gov/33070099/) | The bed nucleus of the stria terminalis and functionally linked neurocircuitry modulate emotion processing and HPA axis dysfunction in posttraumatic stress disorder | GE-EPI | 70.3 | 5/1 | 24m | no data | Axial/axial oblique | Yes | No |
| [**Baczkowski *et al.* 2017**](https://pubmed.ncbi.nlm.nih.gov/28039553/) | Deficient amygdala-prefrontal intrinsic connectivity after effortful emotion regulation in borderline personality disorder | GE-EPI | 27 | 3/0 | 6m | Birdcage;  8-channel | Axial/axial oblique | Yes | No |
| [**Baeken *et al.* 2014**](https://pubmed.ncbi.nlm.nih.gov/24760033/) | Left and right amygdala-mediofrontal cortical functional connectivity is differentially modulated by harm avoidance | SE-EPI | 13 | 4/1 | 5m | 8 channel | Axial/axial oblique* | Yes | No |
| [**Bebko *et al.* 2015**](https://pubmed.ncbi.nlm.nih.gov/25433424/) | Decreased amygdala-insula resting state connectivity in behaviorally and emotionally dysregulated youth | GE-EPI | 31.8 | 3.1/0 | 6m | no data | Axial/axial oblique | Yes | No |
| [**Belleau *et al.* 2020**](https://pubmed.ncbi.nlm.nih.gov/32435666/) | Amygdala functional connectivity in the acute aftermath of trauma prospectively predicts severity of posttraumatic stress symptoms | GE-EPI | 52; 49.2 | 3.7/0; 3.5/0 | 6m; 5m | no data | Sagittal | Yes | No |
| [**Benito-Leon *et al.* 2019**](https://pubmed.ncbi.nlm.nih.gov/31332912/) | Graph theory analysis of resting-state functional magnetic resonance imaging in essential tremor | GE-EPI | 20.4 | 2.8/0 | 6m | 8 channel | Axial/axial oblique | Yes | No |
| [**Ben Simon *et al.* 2017**](https://pubmed.ncbi.nlm.nih.gov/28370703/) | Tired and misconnected: A breakdown of brain modularity following sleep deprivation | GE-EPI | 36 | 4/0 | 6m 50s | Standard | Axial/axial oblique | Yes | No |
| [**Bertolino *et al.* 2005**](https://pubmed.ncbi.nlm.nih.gov/15953488/) | Variation of human amygdala response during threatening stimuli as a function of 5'HTTLPR genotype and personality style | GE-EPI | 70.3 | 5/0 | no data | no data | Axial/axial oblique | Yes | No |
| [**Bickart *et al.* 2012**](https://pubmed.ncbi.nlm.nih.gov/23077058/) | Intrinsic amygdala-cortical functional connectivity predicts social network size in humans | GE-EPI | 27; 8; 48.8 | 3/0; 2/0; 5/0 | no data | 12 channel | Axial/axial oblique* | Yes | Yes |
| [**Bielski *et al.* 2021**](https://pubmed.ncbi.nlm.nih.gov/33338610/) | Parcellation of the human amygdala using recurrence quantification analysis | GE-EPI | 15.6; 15.6; 27 | 2.5/0; 2.5/0; 3/0 | 15m;15m;  12m | 32 channel | Axial/axial oblique* | Yes | Yes |
| [**Bjertrup *et al.* 2022**](https://pubmed.ncbi.nlm.nih.gov/34706300/) | Reduced prefrontal cortex response to own vs. unknown emotional infant faces in mothers with bipolar disorder | GE spiral EPI | 38.7 | 3/0.75 | 6m 18s | 64 channel | Axial/axial oblique* | Yes | No |
| [**Bonduelle *et al.* 2021**](https://pubmed.ncbi.nlm.nih.gov/34295271/) | Exposure to criticism modulates left but not right amygdala functional connectivity in healthy adolescents: Individual influences of perceived and self-criticism | GE-EPI | 27 | 3/0 | no data | 64 channel | Axial/axial oblique | Yes | No |
| [**Bonnet *et al.* 2015**](https://pubmed.ncbi.nlm.nih.gov/26217205/) | The role of the amygdala in the perception of positive emotions: an "intensity detector" | GE-EPI | 18 | 4.5/0 | 15m | no data | Axial/axial oblique | No | No |
| [**Borgers *et al.* 2022**](https://pubmed.ncbi.nlm.nih.gov/35030511/) | Brain functional correlates of emotional face processing in body dysmorphic disorder | GE-EPI | 46.7 | 3.6/0 | no data | no data | Axial/axial oblique | Yes | No |
| [**Brady Jr *et al.* 2016**](https://pubmed.ncbi.nlm.nih.gov/27177299/) | State dependent cortico-amygdala circuit dysfunction in bipolar disorder | GE-EPI | 27 | 3/0 | no data | 12 channel | Axial/axial oblique | Yes | No |
| [**Burklund *et al.* 2007**](https://pubmed.ncbi.nlm.nih.gov/18461157/) | The face of rejection: rejection sensitivity moderates dorsal anterior cingulate activity to disapproving facial expressions | GE-EPI | 29.3 | 3/1 | 5m 30s | no data | Axial/axial oblique | Yes | No |
| [**Cao *et al.* 2022**](https://pubmed.ncbi.nlm.nih.gov/34748823/) | Distinct alterations of amygdala subregional functional connectivity in early-and late-onset obsessive-compulsive disorder | GE-EPI | 70.3 | 5/0 | no data | 8 channel | Axial/axial oblique | Yes | Yes |
| [**Carey *et al.* 2020**](https://pubmed.ncbi.nlm.nih.gov/32979785/) | Anxiety in Parkinson's disease is associated with changes in the brain fear circuit | GE-EPI | 27 | 3/0 | 10m | no data | Axial/axial oblique | Yes | No |
| [**Chen *et al.* 2015**](https://pubmed.ncbi.nlm.nih.gov/25622022/) | Temporal lobe epilepsy: decreased thalamic resting-state functional connectivity and their relationships with alertness performance | GE-EPI | 59.2 | 5/1 | no data | 12 channel | Axial/axial oblique* | Yes | No |
| [**Chen *et al.* 2020**](https://pubmed.ncbi.nlm.nih.gov/32253102/) | Amygdala Functional Connectivity Features in Grief: A Pilot Longitudinal Study | GE-EPI | 56.2 | 4/0 | 8m | Standard transmit and receive | Sagittal | Yes | No |
| [**Cheng *et al.* 2018**](https://pubmed.ncbi.nlm.nih.gov/29767786/) | Functional connectivity of the human amygdala in health and in depression | GE-EPI | 34.7; 40.2 | 3/1; 3.4/0 | 8m | 16 channel;  12 channel | Axial/axial oblique | Yes | Yes |
| [**Cheung *et al.* 2019**](https://pubmed.ncbi.nlm.nih.gov/31708393/) | Uncertainty and surprise jointly predict musical pleasure and amygdala, hippocampus, and auditory cortex activity | GE-EPI | 28.8 | 3.2/0.32 | no data | 32 channel | Axial/axial oblique | No | No |
| [**Chong *et al.* 2017**](https://pubmed.ncbi.nlm.nih.gov/27306407/) | Migraine classification using magnetic resonance imaging resting-state functional connectivity data | GE-EPI^ | 64; 64 | 4/0; 4/0 | 10m | no data | Axial/axial oblique* | Yes | No |
| [**Chumachenko *et al.* 2021**](https://pubmed.ncbi.nlm.nih.gov/33428638/) | Keeping weight off: Mindfulness-Based Stress Reduction alters amygdala functional connectivity during weight loss maintenance in a randomized control trial | GE-EPI | 42.9 | 3.5/0 | 6m 4s | 12 channel | Axial/axial oblique* | Yes | No |
| [**Cisler *et al.* 2015**](https://pubmed.ncbi.nlm.nih.gov/26522869/) | Amygdala response predicts trajectory of symptom reduction during trauma-focused cognitive-behavioral therapy among adolescent girls with PTSD | GE-EPI | 22.5 | 2.5/0.5 | 8m x 2 | 32 channel | Axial/axial oblique | Yes | No |
| [**Cisler *et al.* 2016**](https://pubmed.ncbi.nlm.nih.gov/27524285/) | Changes in functional connectivity of the amygdala during cognitive reappraisal predict symptom reduction during trauma-focused cognitive - behavioral therapy among adolescent girls with post-traumatic stress disorder | GE-EPI | 22.5 | 2.5/0.5 | 6m | 32 channel | Axial/axial oblique | Yes | No |
| [**Cisler 2017**](https://pubmed.ncbi.nlm.nih.gov/28553208/) | Childhood trauma and functional connectivity between amygdala and medial prefrontal cortex: A dynamic functional connectivity and large-scale network perspective | GE-EPI | 27; 22.5 | 3/0; 2.5/0.5 | 7m 30s | 8 channel;  32 channel | Axial/axial oblique | Yes | No |
| [**Cocchi *et al.* 2012**](https://pubmed.ncbi.nlm.nih.gov/23223295/) | Altered functional brain connectivity in a non-clinical sample of young adults with attention-deficit/hyperactivity disorder | GE-EPI | 27 | 3/0 | 8m | no data | Axial/axial oblique | Yes | No |
| [**Connolly *et al.* 2013**](https://pubmed.ncbi.nlm.nih.gov/23910949/) | Resting-state functional connectivity of subgenual anterior cingulate cortex in depressed adolescents | GE-EPI | 27 | 3/0 | 8m 32s | no data | Axial/axial oblique | Yes | No |
| [**Coombs III *et al.* 2014**](https://pubmed.ncbi.nlm.nih.gov/24816735/) | Amygdala perfusion is predicted by its functional connectivity with the ventromedial prefrontal cortex and negative affect | GE-EPI; ASL | 27; 61.8 | 3/0; 5/1.25 | 6m 12s | 12 channel | Axial/axial oblique* | Yes | Yes |
| [**Crane *et al.* 2018**](https://pubmed.ncbi.nlm.nih.gov/30268936/) | Amygdala-orbitofrontal functional connectivity mediates the relationship between sensation seeking and alcohol use among binge-drinking adults | GE-EPI | 36 | 4/0.5 | 10m | 8 channel | Axial/axial oblique* | Yes | No |
| [**Cullen *et al.* 2014**](https://pubmed.ncbi.nlm.nih.gov/25133665/) | Abnormal amygdala resting-state functional connectivity in adolescent depression | GE-EPI | 47.1 | 4/0 | 6m | no data | Axial/axial oblique* | Yes | No |
| [**Delli Pizzi *et al.* 2017a**](https://pubmed.ncbi.nlm.nih.gov/27566606/) | Functional and neurochemical interactions within the amygdala-medial prefrontal cortex circuit and their relevance to emotional processing | GE-EPI | 64.6 | 5/0 | no data | 8 channel | Axial/axial oblique | Yes | No |
| [**Delli Pizzi *et al.* 2017b**](https://pubmed.ncbi.nlm.nih.gov/28386778/) | GABA content within medial prefrontal cortex predicts the variability of fronto-limbic effective connectivity | GE-EPI | 64.6 | 5/0 | no data | 8 channel | Axial/axial oblique | Yes | No |
| [**Demos *et al.* 2008**](https://pubmed.ncbi.nlm.nih.gov/18372291/) | Human amygdala sensitivity to the pupil size of others | GE-EPI | 31.5 | 3.5/0.5 | no data | SENSE head | Axial/axial oblique | Yes | No |
| [**Deng *et al.* 2019**](https://pubmed.ncbi.nlm.nih.gov/31143106/) | Neuroticism modulates the functional connectivity from amygdala to frontal networks in females when avoiding emotional negative pictures | GE-EPI^ | 42.9 | 3.5/0.7 | no data | no data | Axial/axial oblique | Yes | No |
| [**Doerig *et al.* 2016**](https://pubmed.ncbi.nlm.nih.gov/26494872/) | Amygdala response to self-critical stimuli and symptom improvement in psychotherapy for depression | GE-EPI | 30.25 | 4/0 | 12m | 8 channel | Axial/axial oblique | Yes | No |
| [**Dong *et al.* 2020**](https://pubmed.ncbi.nlm.nih.gov/32987837/) | Improvement in uncontrolled eating behavior after laparoscopic sleeve gastrectomy is associated with alterations in the brain-gut-microbiome axis in obese women | GE-EPI^ | 47.3 | 4/0 | 10m | no data | Axial/axial oblique* | Yes | No |
| [**Dufford *et al.* 2019**](https://pubmed.ncbi.nlm.nih.gov/31106452/) | Maternal brain resting-state connectivity in the postpartum period | GE-EPI | 27 | 3/0 | 5m | 32 channel | Axial/axial oblique | Yes | No |
| [**Dutra *et al.* 2017**](https://pubmed.ncbi.nlm.nih.gov/29024194/) | Disrupted cortico-limbic connectivity during reward processing in remitted bipolar I disorder | GE-EPI | 46.2 | 4/0 | 7m | no data | Axial/axial oblique* | Yes | No |
| [**Dvir *et al.* 2020**](https://pubmed.ncbi.nlm.nih.gov/33192645/) | Psychiatric Symptomatology, Mood Regulation, and Resting State Functional Connectivity of the Amygdala: Preliminary Findings in Youth With Mood Disorders and Childhood Trauma | GE-EPI^ | 9.7 | 3/0 | 6m | 8 channel | Axial/axial oblique | Yes | No |
| [**Ely *et al.* 2016**](https://pubmed.ncbi.nlm.nih.gov/26991474/) | Resting-state functional connectivity of the human habenula in healthy individuals: Associations with subclinical depression | GE-EPI^ | 8 | 2/0 | 15m x 4 = 1hr | 32 channel | Axial/axial oblique* | Yes | No |
| [**Fan *et al.* 2014**](https://pubmed.ncbi.nlm.nih.gov/24862297/) | Early life stress modulates amygdala-prefrontal functional connectivity: Implications for oxytocin effects | GE-EPI | 27 | 3/0 | no data | 12 channel | Axial/axial oblique | Yes | No |
| [**Fan *et al.* 2015**](https://pubmed.ncbi.nlm.nih.gov/25924202/) | Amygdala-hippocampal connectivity changes during acute psychosocial stress: joint effect of early life stress and oxytocin | GE-EPI | 27 | 3/0 | 7m x 3 | 12 channel | Axial/axial oblique | Yes | No |
| [**Fan *et al.* 2020**](https://pubmed.ncbi.nlm.nih.gov/31628591/) | Altered functional connectivity of the amygdala in Crohn's disease | GE-EPI | 70.3 | 5/0 | no data | no data | Axial/axial oblique* | Yes | No |
| [**Fateh *et al.* 2020**](https://pubmed.ncbi.nlm.nih.gov/32738725/) | Disrupted dynamic functional connectivity in right amygdalar subregions differentiates bipolar disorder from major depressive disorder | GE-EPI | 45 | 3.2/0 | no data | ‘8-channel proto-type quadrature birdcage head’ | Axial/axial oblique* | Yes | Yes |
| [**Feng *et al.* 2014**](https://pubmed.ncbi.nlm.nih.gov/24194579/) | Memory consolidation of fear conditioning: bi-stable amygdala connectivity with dorsal anterior cingulate and medial prefrontal cortex | GE-EPI | 27 | 3/0.99 | no data | no data | Axial/axial oblique* | Yes | No |
| [**Fernández-Alcántara *et al.*  2020**](https://pubmed.ncbi.nlm.nih.gov/32245009/) | Increased amygdala activations during the emotional experience of death-related pictures in complicated grief: an fMRI study | GE-EPI | 27 | 3/1 | no data | 32 channel | Axial/axial oblique | Yes | No |
| [**Feurer *et al.* 2021**](https://pubmed.ncbi.nlm.nih.gov/33621397/) | Resting state functional connectivity correlates of rumination and worry in internalizing psychopathologies | GE-EPI | 35.4 | 3/0 | no data | 8 channel | Axial/axial oblique | Yes | No |
| [**Fisher *et al.* 2017**](https://pubmed.ncbi.nlm.nih.gov/27649641/) | Pharmacologically induced sex hormone fluctuation effects on resting-state functional connectivity in a risk model for depression: a randomized trial | GE-EPI | 27 | 3/0 | 10m | 32 channel | Axial/axial oblique* | Yes | No |
| [**Fitzgerald *et al.* 2019**](https://pubmed.ncbi.nlm.nih.gov/30408261/) | Transdiagnostic neural correlates of volitional emotion regulation in anxiety and depression | GE-EPI^ | 35.4 | 3/0 | no data | Standard | Axial/axial oblique* | Yes | No |
| [**Frijling *et al.* 2016**](https://pubmed.ncbi.nlm.nih.gov/26346640/) | Intranasal Oxytocin Affects Amygdala Functional Connectivity after Trauma Script-Driven Imagery in Distressed Recently Trauma-Exposed Individuals | GE-EPI | 15.7 | 3/0 | 8m | 16 channel | Axial/axial oblique | Yes | No |
| [**Fu *et al.* 2015**](https://pubmed.ncbi.nlm.nih.gov/25880400/) | Multimodal functional and structural neuroimaging investigation of major depressive disorder following treatment with duloxetine | GE-EPI | 42.2 | 3/0.3 | 8m 30s | no data | Axial/axial oblique | Yes | No |
| [**Gamer *et al.* 2009**](https://pubmed.ncbi.nlm.nih.gov/19605649/) | Amygdala activation predicts gaze toward fearful eyes | GE-EPI | 8 | 2/1 | no data | 12 channel | Axial/axial oblique | Yes | No |
| [**Gamer *et al.* 2010**](https://pubmed.ncbi.nlm.nih.gov/20421469/) | Different amygdala subregions mediate valence-related and attentional effects of oxytocin in humans | GE-EPI | 8 | 2/0 | no data | 32 channel | Axial/axial oblique | No | Yes |
| [**Ganella *et al.* 2017**](https://pubmed.ncbi.nlm.nih.gov/29255411/) | Prefrontal-amygdala connectivity and state anxiety during fear extinction recall in adolescents | GE-EPI | 27 | 3/0 | no data | 32 channel | Axial/axial oblique* | Yes | No |
| [**Gao *et al.* 2016**](https://pubmed.ncbi.nlm.nih.gov/27994546/) | Decreased subcortical and increased cortical degree centrality in a nonclinical college student sample with subclinical depressive symptoms: a resting-state fMRI study | GE-EPI | 34.2 | 3.5/0 | no data | 8 channel | Axial/axial oblique* | Yes | No |
| [**Ge *et al.* 2021**](https://pubmed.ncbi.nlm.nih.gov/34207133/) | Aerobic Exercise Decreases Negative Affect by Modulating Orbitofrontal-Amygdala Connectivity in Adolescents | GE-EPI | 46.2 | 4/0 | no data | 12 channel | Axial/axial oblique | Yes | No |
| [**Geng *et al.* 2016**](https://pubmed.ncbi.nlm.nih.gov/26834594/) | Decreased intra-and inter-salience network functional connectivity is related to trait anxiety in adolescents | GE-EPI | 35.4 | 3/1 | 7m 30s | no data | Axial/axial oblique | Yes | Yes |
| [**Gilam *et al.* 2017**](https://pubmed.ncbi.nlm.nih.gov/29326568/) | Tracing the neural carryover effects of interpersonal anger on resting-state fMRI in men and their relation to traumatic stress symptoms in a subsample of soldiers | GE-EPI | 7.3 | 3/0 | 6m | 8 channel | Axial/axial oblique | Yes | No |
| [**Gingnell *et al.* 2013**](https://pubmed.ncbi.nlm.nih.gov/23219471/) | Oral contraceptive use changes brain activity and mood in women with previous negative affect on the pill - A double-blinded, placebo-controlled randomized trial of a levonorgestrel-containing combined oral contraceptive | GE-EPI | 27 | 3/0 | no data | 8 channel | Axial/axial oblique* | Yes | No |
| [**Gorka *et al.* 2016**](https://pubmed.ncbi.nlm.nih.gov/26647971/) | Cannabinoid Modulation of Frontolimbic Activation and Connectivity during Volitional Regulation of Negative Affect | GE reverse spiral;  GE-EPI | 35.4 | 3/0; 3/0 | no data | Standard;  8 channel | Axial/axial oblique | Yes | No |
| [**Green *et al.* 2016**](https://pubmed.ncbi.nlm.nih.gov/27343889/) | Salience Network Connectivity in Autism Is Related to Brain and Behavioral Markers of Sensory Overresponsivity | GE-EPI | 27/29.3 | 3/0; 3/0 | 6m; 5m 43s | no data | Axial/axial oblique* | Yes | No |
| [**Hagland *et al.* 2021**](https://pubmed.ncbi.nlm.nih.gov/33841194/) | Disentangling Within- and Between-Person Effects During Response Inhibition in Obsessive-Compulsive Disorder | GE-EPI^ | 33.1 | 2.8/0.2 | no data | 8 channel | Axial/axial oblique* | Yes | No |
| [**Han *et al.* 2021**](https://pubmed.ncbi.nlm.nih.gov/32720182/) | Self-reported experiences of discrimination in older black adults are associated with insula functional connectivity | GE-EPI | 36.9 | 3.3/0 | 8m18s | no data | Axial/axial oblique | Yes | No |
| [**Hansen *et al.* 2020**](https://pubmed.ncbi.nlm.nih.gov/33075046/) | Adults vs. neonates: Differentiation of functional connectivity between the basolateral amygdala and occipitotemporal cortex | GE-EPI; GE-EPI | 8; 9.9 | 2/0; 2.15/0 | 15m; 15m | 32 channel;  32 channel | Axial/axial oblique* | Yes | Yes |
| [**Hardee *et al.* 2008**](https://pubmed.ncbi.nlm.nih.gov/19015094/) | The left amygdala knows fear: laterality in the amygdala response to fearful eyes | GE spiral in-out | 14.1 | 4/1 | no data | no data | Axial/axial oblique | Yes | No |
| [**Herringa *et al.* 2013**](https://pubmed.ncbi.nlm.nih.gov/24191026/) | Childhood maltreatment is associated with altered fear circuitry and increased internalizing symptoms by late adolescence | GE-EPI | 61.3 | 5/0 | 7m | 8 channel | Sagittal | Yes | No |
| [**Ho *et al.* 2014**](https://pubmed.ncbi.nlm.nih.gov/24268546/) | Functional connectivity of negative emotional processing in adolescent depression | GE-EPI | 33.6 | 2.6/1.4 | no data | 8 channel | Axial/axial oblique | Yes | No |
| [**Hofmann *et al.* 2019**](https://pubmed.ncbi.nlm.nih.gov/30829454/) | Resting-state fMRI effective connectivity between the bed nucleus of the stria terminalis and amygdala nuclei | GE-EPI | 8 | 2/0 | 15m x 4 = 1hr | 32 channel | Axial/axial oblique | Yes | Yes |
| [**Horne *et al.* 2018**](https://pubmed.ncbi.nlm.nih.gov/29339309/) | Late chronotype is associated with enhanced amygdala reactivity and reduced fronto-limbic functional connectivity to fearful versus happy facial expressions | GE-EPI | 27 | 3/0 | 5m 40s | 32 channel | Axial/axial oblique* | No | No |
| [**Hsu *et al.* 2015**](https://pubmed.ncbi.nlm.nih.gov/25671315/) | The magical activation of left amygdala when reading Harry Potter: an fMRI study on how descriptions of supra-natural events entertain and enchant | GE EPI | 27 | 3/0 | no data | no data | Axial/axial oblique* | Yes | No |
| [**Jacobs *et al.* 2016**](https://pubmed.ncbi.nlm.nih.gov/26784396/) | Decoupling of the amygdala to other salience network regions in adolescent-onset recurrent major depressive disorder | GE SS reverse spiral;  GE-EPI | 39.1; 35.4 | 4/0; 3/0 | no data | no data | Axial/axial oblique* | Yes | No |
| [**Jeong *et al.* 2019**](https://pubmed.ncbi.nlm.nih.gov/30477594/) | Altered functional connectivity in the fear network of firefighters with repeated traumatic stress | GE-EPI | 40.6 | 3.44/0 | no data | no data | Axial/axial oblique* | Yes | No |
| [**Jiang *et al.* 2021**](https://pubmed.ncbi.nlm.nih.gov/33359350/) | Intrinsic, dynamic and effective connectivity among large-scale brain networks modulated by oxytocin | GE-EPI | 27 | 3/0 | 8m 30s | no data | Axial/axial oblique* | Yes | Yes |
| [**Kaag *et al.* 2018**](https://pubmed.ncbi.nlm.nih.gov/29593581/) | Enhanced amygdala-striatal functional connectivity during the processing of cocaine cues in male cocaine users with a history of childhood trauma | GE-EPI | 27 | 3/3 | no data | 32 channel | Axial/axial oblique | Yes | No |
| [**Kalmar *et al.* 2009**](https://pubmed.ncbi.nlm.nih.gov/19454919/) | Relation between amygdala structure and function in adolescents with bipolar disorder | GE-EPI | 42.2 | 3/0 | no data | no data | Axial/axial oblique | Yes | No |
| [**Kanel *et al.* 2022**](https://pubmed.ncbi.nlm.nih.gov/35178519/) | Neonatal amygdala resting-state functional connectivity and socio-emotional development in very preterm children | GE-EPI | 25 | 4/0 | 6m 24s | 8 channel | Axial/axial oblique* | Yes | No |
| [**Ke *et al.* 2017**](https://pubmed.ncbi.nlm.nih.gov/27722829/) | Post-traumatic stress influences local and remote functional connectivity: a resting-state functional magnetic resonance imaging study | GE-EPI | 46.5 | 3.6/0 | no data | ‘Standard’ | Axial/axial oblique | Yes | No |
| [**Kemmotsu *et al.* 2013**](https://pubmed.ncbi.nlm.nih.gov/24176688/) | Alterations in functional connectivity between the hippocampus and prefrontal cortex as a correlate of depressive symptoms in temporal lobe epilepsy | GE-EPI | 29.5 | 2.5/0 | no data | 8 channel | Axial/axial oblique | Yes | No |
| [**Kemmotsu *et al.* 2014**](https://pubmed.ncbi.nlm.nih.gov/25223729/) | Frontolimbic brain networks predict depressive symptoms in temporal lobe epilepsy | GE-EPI | 29.5 | 2.5/0 | no data | 8 channel | Axial/axial oblique* | Yes | No |
| [**King *et al.* 2016a**](https://pubmed.ncbi.nlm.nih.gov/27703434/) | A pilot study of mindfulness-based exposure therapy in OEF/OIF combat veterans with PTSD: altered medial frontal cortex and amygdala responses in socialâ€“emotional processing | GE-EPI | 35.4 | 3/0 | no data | no data | Axial/axial oblique* | Yes | No |
| [**King *et al.* 2016b**](https://pubmed.ncbi.nlm.nih.gov/27038410/) | Altered default mode network (DMN) resting state functional connectivity following a mindfulness-based exposure therapy for posttraumatic stress disorder (PTSD) in combat veterans of Afghanistan and Iraq | GE-EPI | 35.4 | 3/0 | no data | no data | Axial/axial oblique | Yes | No |
| [**Kliemann *et al.* 2012**](https://pubmed.ncbi.nlm.nih.gov/22787032/) | The role of the amygdala in atypical gaze on emotional faces in autism spectrum disorders | GE-EPI | 8 | 2/0 | no data | 12 channel | Axial/axial oblique | No | No |
| [**Klumpp *et al.* 2018**](https://pubmed.ncbi.nlm.nih.gov/29896128/) | Self-reported sleep quality modulates amygdala resting-state functional connectivity in anxiety and depression | GE-EPI | 35.4 | 3/0 | no data | 8 channel | Axial/axial oblique | Yes | No |
| [**Ko *et al.* 2015**](https://pubmed.ncbi.nlm.nih.gov/25448779/) | Altered gray matter density and disrupted functional connectivity of the amygdala in adults with Internet gaming disorder | GE-EPI | 56.3 | 4/0 | no data | no data | Axial/axial oblique | Yes | No |
| [**Korgaonkar *et al.* 2019**](https://pubmed.ncbi.nlm.nih.gov/30343134/) | Amygdala Activation and Connectivity to Emotional Processing Distinguishes Asymptomatic Patients With Bipolar Disorders and Unipolar Depression | GE-EPI | 49.2 | 3.5/0 | 5m 8s | 8 channel | Axial/axial oblique | Yes | No |
| **Korgaonkar *et al.* 2021** | Neural correlates of emotional processing in panic disorder | GE-EPI | 49.2 | 3.5/0 | no data | 8 channel | Axial/axial oblique | Yes | No |
| [**Kramer *et al.* 2015**](https://pubmed.ncbi.nlm.nih.gov/26314945/) | Dynamic Amygdala Influences on the Fronto-Striatal Brain Mechanisms Involved in Self-Control of Impulsive Desires | GE-EPI | 27 | 3/0.6 | no data | 8 channel | Axial/axial oblique | Yes | No |
| [**Krause-Utz *et al.* 2018**](https://pubmed.ncbi.nlm.nih.gov/28526931/) | Reduced amygdala reactivity and impaired working memory during dissociation in borderline personality disorder | GE-EPI | 27 | 3/0 | no data | no data | Axial/axial oblique | Yes | No |
| [**Kumar *et al.* 2015**](https://www.ncbi.nlm.nih.gov/pmc/articles/PMC4376540/) | Oxytocin Affects the Connectivity of the Precuneus and the Amygdala: A Randomized, Double-Blinded, Placebo-Controlled Neuroimaging Trial | GE-EPI | 31.5 | 3.5/0 | 5m | 32 channel | Axial/axial oblique | Yes | No |
| [**Labrenz *et al.* 2019**](https://pubmed.ncbi.nlm.nih.gov/30243957/) | Altered temporal variance and functional connectivity of BOLD signal is associated with state anxiety during acute systemic inflammation | GE-EPI | 23.5 | 3/0.6 | 10m | 32 channel | Axial/axial oblique | Yes | No |
| [**Laeger *et al.* 2014**](https://pubmed.ncbi.nlm.nih.gov/25396729/) | Of 'disgrace' and 'pain' - Corticolimbic interaction patterns for disorder-relevant and emotional words in social phobia | GE-EPI | 46.5 | 3.6/0 | 6m 40s | ‘Circularly Polarized Transmit and Receive birdcage’ | Axial/axial oblique | Yes | No |
| [**Lee *et al.* 2013**](https://pubmed.ncbi.nlm.nih.gov/23273106/) | Amygdala activity contributes to the dissociative effect of cannabis on pain perception | GE-EPI | 27 | 3/0 | 5m; 15m | ‘Transmit and receive birdcage’ | Axial/axial oblique* | Yes | No |
| [**Lee *et al.* 2020**](https://pubmed.ncbi.nlm.nih.gov/33138873/) | Aberrant functional connectivity of neural circuits associated with thought-action fusion in patients with obsessive-compulsive disorder | GE-EPI | 51.7 | 4/0 | 14m 56s | 24 channel | Axial/axial oblique* | Yes | No |
| [**Leicht *et al.* 2013**](https://pubmed.ncbi.nlm.nih.gov/23059050/) | Benzodiazepines counteract rostral anterior cingulate cortex activation induced by cholecystokinin-tetrapeptide in humans | GE-EPI | 27 | 3/0.75 | 15m | no data | Axial/axial oblique* | Yes | No |
| [**Leutgeb *et al.* 2016**](https://pubmed.ncbi.nlm.nih.gov/26523791/) | Altered cerebellar-amygdala connectivity in violent offenders: A resting-state fMRI study | GE-EPI^ | 42.9 | 3.5/0 | 8m 1s | 32 channel | Axial/axial oblique* | Yes | No |
| [**Li  *et al.* 2015**](https://pubmed.ncbi.nlm.nih.gov/26424431/) | Amygdala network dysfunction in late-life depression phenotypes: Relationships with symptom dimensions | GE-EPI | 56.25 | 4/0 | 8m | ‘Standard transmit and receive quadrature’ | Axial/axial oblique* | Yes | No |
| [**Li *et al.* 2016**](https://pubmed.ncbi.nlm.nih.gov/27867352/) | Aberrant functional connectivity between the amygdala and the temporal pole in drug-free generalized anxiety disorder | GE-EPI | 70.3 | 5/0 | 2m 44s; 5m 14s | 12 channel | Axial/axial oblique | Yes | No |
| [**Li *et al.* 2019**](https://pubmed.ncbi.nlm.nih.gov/31824159/) | Resting-state functional MRI study: Connection strength of brain networks in DR patients | GE-EPI^ | 64 | 4/0 | 8m | 8 channel | Axial/axial oblique* | Yes | No |
| [**Lips *et al.* 2014**](https://pubmed.ncbi.nlm.nih.gov/24965310/) | Resting-state functional connectivity of brain regions involved in cognitive control, motivation, and reward is enhanced in obese females | GE-EPI | 20.8 | 2.75/0.25 | no data | 8 channel | Axial/axial oblique | Yes | No |
| [**Liu *et al.* 2021**](https://pubmed.ncbi.nlm.nih.gov/33205889/) | Altered functional connectivity of the amygdala and its subregions in typhoon-related post-traumatic stress disorder | GE-EPI | 57.6 | 3.6/0 | 8m 28s | 32 channel | Axial/axial oblique | Yes | Yes |
| [**Loewenstern *et al.* 2019**](https://pubmed.ncbi.nlm.nih.gov/30711709/) | Interactive effect of 5-HTTLPR and BDNF polymorphisms on amygdala intrinsic functional connectivity and anxiety | GE-EPI | 27 | 3/0 | 5m 4 sec | no data | Axial/axial oblique* | Yes | No |
| [**Lu *et al.* 2019**](https://pubmed.ncbi.nlm.nih.gov/30633429/) | Connectome-based model predicts individual differences in propensity to trust | GE-EPI | 42.9 | 3.5/0.7 | 5m | no data | Axial/axial oblique | Yes | No |
| [**Lu *et al.* 2021**](https://pubmed.ncbi.nlm.nih.gov/34580419/) | Acute neurofunctional effects of escitalopram during emotional processing in pediatric anxiety: a double-blind, placebo-controlled trial | GE EPI | 23.3 | 3/0 | 8m 38s | 32 channel | Axial/axial oblique | Yes | No |
| [**Ma *et al.* 2021**](https://www.ncbi.nlm.nih.gov/pmc/articles/PMC8968690/) | Resting-state directional connectivity and anxiety and depression symptoms in adult cannabis users | GE-EPI; GE-EPI | 8; 27 | 2/0; 3/0 | 7m 12s; 6m | 32 channel;  32 channel | Axial/axial oblique* | Yes | No |
| [**Man *et al.* 2019**](https://pubmed.ncbi.nlm.nih.gov/30423467/) | Altered amygdala circuits underlying valence processing among manic and depressed phases in bipolar adults | GE-EPI | 46.24 | 4/0 | 8m 44s | no data | Axial/axial oblique* | Yes | No |
| [**Mangia *et al.* 2017**](https://www.ncbi.nlm.nih.gov/pmc/articles/PMC5742124/) | Multi-modal Brain MRI in Subjects with PD and iRBD | GE-EPI | 27 | 3/0 | no data | 32 channel | Axial/axial oblique | Yes | No |
| [**Mansson *et al.* 2013**](https://pubmed.ncbi.nlm.nih.gov/24064198/) | Altered neural correlates of affective processing after internet-delivered cognitive behavior therapy for social anxiety disorder | GE-EPI | 23.1 | 3.4/0 | no data | 32 channel | Axial/axial oblique* | Yes | No |
| [**Mareckova *et al.* 2016**](https://www.ncbi.nlm.nih.gov/pmc/articles/PMC5053909/) | Brain activity and connectivity in response to negative affective stimuli: Impact of dysphoric mood and sex across diagnoses | SE -PI | 48.8 | 5/0 | no data | 12 channel | Axial/axial oblique | Yes | No |
| [**McLeod *et al.* 2014**](https://pubmed.ncbi.nlm.nih.gov/24818082/) | Functional connectivity of neural motor networks is disrupted in children with developmental coordination disorder and attention-deficit/hyperactivity disorder | GE-EPI | 47.3 | 4/0 | 5m | 8 channel | Axial/axial oblique* | Yes | No |
| [**Mertens *et al.* 2020**](https://pubmed.ncbi.nlm.nih.gov/31941394/) | Therapeutic mechanisms of psilocybin: Changes in amygdala and prefrontal functional connectivity during emotional processing after psilocybin for treatment-resistant depression | GE-EPI | 27 | 3/0 | 8m | 12 channel | Axial/axial oblique | Yes | No |
| [**Michely *et al.* 2020**](https://pubmed.ncbi.nlm.nih.gov/31542358/) | Distinct processing of aversive experience in amygdala subregions | GE-EPI | 27 | 3/0 | no data | 32 channel | Axial/axial oblique | Yes | Yes |
| [**Minzenberg *et al.* 2007**](https://pubmed.ncbi.nlm.nih.gov/17601709/) | Fronto-limbic dysfunction in response to facial emotion in borderline personality disorder: an event-related fMRI study | GE-EPI | 26.9 | 2.5/0.825 | 4m 48s | no data | Axial/axial oblique | Yes | No |
| [**Miskowiak *et al.* 2017**](https://pubmed.ncbi.nlm.nih.gov/28351201/) | Does a single session of electroconvulsive therapy alter the neural response to emotional faces in depression? A randomised sham-controlled functional magnetic resonance imaging study | GE spiral EPI | 27 | 3/0 | 5m 28s | 8 channel | Axial/axial oblique* | Yes | No |
| [**Monk *et al.* 2008**](https://pubmed.ncbi.nlm.nih.gov/17986682/) | Amygdala and nucleus accumbens activation to emotional facial expressions in children and adolescents at risk for major depression | GE-EPI | 46.4 | 3.3/0 | no data | no data | Axial/axial oblique | Yes | No |
| [**Motomura *et al.* 2013**](https://pubmed.ncbi.nlm.nih.gov/23418586/) | Sleep debt elicits negative emotional reaction through diminished amygdala-anterior cingulate functional connectivity | GE-EPI | 36 | 4/1 | no data | no data | Axial/axial oblique | Yes | No |
| [**Motomura *et al.* 2017**](https://pubmed.ncbi.nlm.nih.gov/28977527/) | Two days' sleep debt causes mood decline during resting state via diminished amygdala-prefrontal connectivity | GE-EPI | 39.3 | 3.4/4.25 | no data | no data | Axial/axial oblique | Yes | No |
| [**Muehlhan *et al.* 2015**](https://pubmed.ncbi.nlm.nih.gov/26303978/) | Epigenetic variation in the serotonin transporter gene predicts resting state functional connectivity strength within the salience-network | GE-EPI | 35.4 | 3/0.75 | no data | 12 channel | Axial/axial oblique | Yes | No |
| [**Nakataki *et al.* 2017**](https://pubmed.ncbi.nlm.nih.gov/27644128/) | Glucocorticoid Administration Improves Aberrant Fear-Processing Networks in Spider Phobia | GE-EPI | 38.7 | 3/0 | no data | 12 channel | Axial/axial oblique | Yes | No |
| [**Nicholson *et al.* 2018**](https://pubmed.ncbi.nlm.nih.gov/30004602/) | Intrinsic connectivity network dynamics in PTSD during amygdala downregulation using real-time fMRI neurofeedback: A preliminary analysis | GE-EPI | 27 | 3/1 | no data | 32 channel | Axial/axial oblique | Yes | No |
| [**Oathes *et al.* 2015**](https://pubmed.ncbi.nlm.nih.gov/25444162/) | Neurobiological signatures of anxiety and depression in resting-state functional magnetic resonance imaging | GE spiral in/out | 47.3 | 4/0.5 | 8 m | 8 channel | Axial/axial oblique | Yes | No |
| [**Ochsner *et al.* 2004**](https://pubmed.ncbi.nlm.nih.gov/15488398/) | For better or for worse: neural systems supporting the cognitive down-and up-regulation of negative emotion | GE spiral in/out | 56.3 | 4/1 | no data | no data | Axial/axial oblique | Yes | No |
| [**Pahapill *et al.* 2020**](https://pubmed.ncbi.nlm.nih.gov/32074111/) | Functional connectivity and structural analysis of trial spinal cord stimulation responders in failed back surgery syndrome | GE-EPI | 56.3 | 4/0 | 6m | ‘Standard quadrature transmit and receive’ | Sagittal | Yes | No |
| [**Pando-Naude *et al.* 2019**](https://pubmed.ncbi.nlm.nih.gov/31664132/) | Functional connectivity of music-induced analgesia in fibromyalgia | GE-EPI | 12 | 3/0 | 5m | 32 channel | Axial/axial oblique | Yes | No |
| [**Pang *et al.* 2016**](https://www.ncbi.nlm.nih.gov/pmc/articles/PMC5073227/) | Extraversion and neuroticism related to the resting-state effective connectivity of amygdala | GE-EPI | 61.9 | 4.4/0 | no data | no data | Axial/axial oblique | Yes | No |
| [**Panikratova *et al.* 2020**](https://pubmed.ncbi.nlm.nih.gov/32342318/) | Functional Connectomic Approach to Studying Selank and Semax Effects | GE-EPI | 13 | 4/0 | 5m | no data | Axial/axial oblique* | Yes | No |
| [**Perlman *et al.* 2012**](https://pubmed.ncbi.nlm.nih.gov/22401827/) | Amygdala response and functional connectivity during emotion regulation: a study of 14 depressed adolescents | GE-EPI | 33.6 | 2.6/1.4 | 9m 36s | 8 channel | Axial/axial oblique | Yes | No |
| [**Prather *et al.* 2013**](https://www.ncbi.nlm.nih.gov/pubmed/23592753) | Impact of sleep quality on amygdala reactivity, negative affect, and perceived stress | 'Inverse spiral' | 56.3 | 4/0 | 6m 30s | 8 channel | Axial/axial oblique | Yes | No |
| [**Qiao *et al.* 2020**](https://pubmed.ncbi.nlm.nih.gov/32871697/) | Brain functional abnormalities in the amygdala subregions is associated with anxious depression | GE-EPI | 56.3 | 4/0 | 6m 45s | no data | Axial/axial oblique* | Yes | Yes |
| [**Qin *et al.* 2014**](https://pubmed.ncbi.nlm.nih.gov/24268662/) | Amygdala subregional structure and intrinsic functional connectivity predicts individual differences in anxiety during early childhood | GE spiral in/spiral out | 39.1 | 4/0.5 | 8m | ‘Custom built’ | Axial/axial oblique | Yes | Yes |
| [**Quinque *et al.* 2014**](https://pubmed.ncbi.nlm.nih.gov/24636515/) | Structural and functional MRI study of the brain, cognition and mood in long-term adequately treated Hashimoto's thyroiditis | GE-EPI | 36 | 4/0 | no data | 32 channel | Axial/axial oblique* | Yes | No |
| [**Rabany *et al.* 2017**](https://www.ncbi.nlm.nih.gov/pubmed/28478685) | Resting-state functional connectivity in generalized anxiety disorder and social anxiety disorder: evidence for a dimensional approach | GE-EPI | 57.8 | 5/0 | 5m 15s | no data | Axial/axial oblique* | Yes | No |
| [**Rausch *et al.* 2016**](https://www.ncbi.nlm.nih.gov/pubmed/26823966) | Altered functional connectivity of the amygdaloid input nuclei in adolescents and young adults with autism spectrum disorder: a resting state fMRI study | DE GE-EPI | 10 | 2.5/0 | 16m | 32 channel | Axial/axial oblique* | Yes | Yes |
| [**Reggente *et al.* 2018**](https://www.ncbi.nlm.nih.gov/pmc/articles/PMC5834692/) | Multivariate resting-state functional connectivity predicts response to cognitive behavioral therapy in obsessive-compulsive disorder | GE EPI | 27 | 3/1 | 7m | 12 channel | Axial/axial oblique | Yes | No |
| [**Reich *et al.* 2019**](https://pubmed.ncbi.nlm.nih.gov/31553999/) | Amygdala Resting State Connectivity Differences between Bipolar II and Borderline Personality Disorders | GE EPI | 42.9 | 3.5/0 | 6m 47s | 32 channel | Coronal/coronal oblique | Yes | No |
| [**Rey *et al.* 2016**](https://pubmed.ncbi.nlm.nih.gov/26945530/) | Resting-state functional connectivity of emotion regulation networks in euthymic and non-euthymic bipolar disorder patients | GE EPI | 32.8 | 3.2/0.8 | no data | 32 channel | Axial/axial oblique | Yes | No |
| [**Rich *et al.* 2008**](https://pubmed.ncbi.nlm.nih.gov/18181882/) | Neural connectivity in children with bipolar disorder: Impairment in the face emotion processing circuit | GE-EPI | 70.3 | 5/0 | no data | no data | Axial/axial oblique | Yes | No |
| [**Riedel *et al.* 2019**](https://pubmed.ncbi.nlm.nih.gov/31268615/) | Modulating functional connectivity between medial frontopolar cortex and amygdala by inhibitory and excitatory transcranial magnetic stimulation | GE-EPI | 18.8 | 3/0 | no data | 32 channel | Axial/axial oblique* | Yes | No |
| [**Rogers *et al.* 2017**](https://pubmed.ncbi.nlm.nih.gov/28117062/) | Neonatal Amygdala Functional Connectivity at Rest in Healthy and Preterm Infants and Early Internalizing Symptoms | GE-EPI | 13.4 | 2.4/0 | 9m 36s | ‘Infant-specific quadrature head’ | Axial/axial oblique | Yes | No |
| [**Rohr *et al.* 2016**](https://pubmed.ncbi.nlm.nih.gov/26991156/) | The neural networks of subjectively evaluated emotional conflicts | GE-EPI | 36 | 4/0 | no data | no data | Axial/axial oblique* | Yes | No |
| [**Roy *et al.* 2013**](https://pubmed.ncbi.nlm.nih.gov/23452685/) | Intrinsic functional connectivity of amygdala-based networks in adolescent generalized anxiety disorder | GE-EPI | 36 | 4/0 | 6m | no data | Axial/axial oblique* | Yes | Yes |
| [**Rzepa *et al.* 2017**](https://pubmed.ncbi.nlm.nih.gov/28340244/) | Bupropion Administration Increases Resting-State Functional Connectivity in Dorso-Medial Prefrontal Cortex | GE-EPI | 13.8 | 2.4/0 | no data | 32 channel | Axial/axial oblique | Yes | No |
| [**Salzwedel *et al.* 2019**](https://pubmed.ncbi.nlm.nih.gov/30316743/) | Development of amygdala functional connectivity during infancy and its relationship with 4-year behavioral outcomes | GE-EPI | 64 | 4/0 | no data | ‘Circular Polarization’ | Axial/axial oblique* | Yes | No |
| [**Sanz-Arigita *et al.* 2021**](https://pubmed.ncbi.nlm.nih.gov/33772591/) | Brain reactivity to humorous films is affected by insomnia | GE-EPI | 15.6 | 2.5/0 | no data | 64 channel | Axial/axial oblique* | Yes | No |
| [**Satterthwaite *et al.* 2010**](https://pubmed.ncbi.nlm.nih.gov/20194482/) | Association of enhanced limbic response to threat with decreased cortical facial recognition memory response in schizophrenia | GE-EPI | 35.4 | 3/0 | no data | no data | Axial/axial oblique | Yes | No |
| [**Satyshur *et al.* 2018**](https://pubmed.ncbi.nlm.nih.gov/29949111/) | Functional connectivity of reflective and brooding rumination in depressed and healthy women | GE-EPI | 8.7 | 3/0 | 10m | 32 channel | Axial/axial oblique | Yes | No |
| [**Savic *et al.* 2017**](https://pubmed.ncbi.nlm.nih.gov/28070912/) | Role of testosterone and Y chromosome genes for the masculinization of the human brain | GE-EPI | 15.2 | 3/0 | 8m | 32 channel | Axial/axial oblique | Yes | No |
| [**Schmitt *et al.* 2020**](https://pubmed.ncbi.nlm.nih.gov/32300362/) | Affective Modulation after High-Intensity Exercise Is Associated with Prolonged Amygdalar-Insular Functional Connectivity Increase | GE-EPI | 46.4 | 3.59/0 | 11m | 8 channel | Axial/axial oblique | Yes | No |
| [**Seo *et al.* 2019**](https://pubmed.ncbi.nlm.nih.gov/31455572/) | In Trauma-Exposed Individuals, Self-reported Hyperarousal and Sleep Architecture Predict Resting-State Functional Connectivity in Frontocortical and Paralimbic Regions. | GE-EPI | 22.5 | 2.5/0 | 10m | 32 channel | Axial/axial oblique* | Yes | No |
| [**Shou *et al.* 2017**](https://pubmed.ncbi.nlm.nih.gov/28275546/) | Cognitive behavioral therapy increases amygdala connectivity with the cognitive control network in both MDD and PTSD | 'gradient spin echo' | 64 | 4/0 | 7m 42s | no data | Axial/axial oblique* | Yes | No |
| [**Sladky *et al.* 2015**](https://pubmed.ncbi.nlm.nih.gov/25536499/) | (S)-citalopram influences amygdala modulation in healthy subjects: a randomized placebo-controlled double-blind fMRI study using dynamic causal modeling | GE-EPI | 9.7 | 3/0.5 | no data | no data | Axial/axial oblique | No | No |
| [**Soe *et al.* 2018**](https://pubmed.ncbi.nlm.nih.gov/29094774/) | Perinatal maternal depressive symptoms alter amygdala functional connectivity in girls | GE-EPI | 27 | 3/0 | 5m 27s; 3m 19s | 32 channel | Axial/axial oblique | Yes | No |
| [**Song *et al.* 2015**](https://pubmed.ncbi.nlm.nih.gov/25762915/) | Love-related changes in the brain: a resting-state functional magnetic resonance imaging study | GE-EPI | 35.4 | 3/0 | 8m 4s | no data | Axial/axial oblique* | Yes | No |
| [**Sripada *et al.* 2014**](https://pubmed.ncbi.nlm.nih.gov/24302681/) | The neurosteroids allopregnanolone and dehydroepiandrosterone modulate resting-state amygdala connectivity | GE-EPI | 35.4 | 3/0 | 8m | no data | Axial/axial oblique* | Yes | No |
| [**Steffens *et al.* 2017**](https://pubmed.ncbi.nlm.nih.gov/28457805/) | Negative Affectivity, Aging, and Depression: Results From the Neurobiology of Late-Life Depression (NBOLD) Study | GE-EPI | 27 | 3/0 | 5m | 32 channel | Axial/axial oblique | Yes | No |
| [**Struck *et al.* 2021**](https://pubmed.ncbi.nlm.nih.gov/33611101/) | Regional and global resting-state functional MR connectivity in temporal lobe epilepsy: Results from the Epilepsy Connectome Project | GE-EPI | 8 | 2/0 | no data | 32 channel | Axial/axial oblique* | Yes | No |
| [**Tei *et al.* 2020**](https://pubmed.ncbi.nlm.nih.gov/32041879/) | Brain and behavioral alterations in subjects with social anxiety dominated by empathic embarrassment | GE-EPI | 27 | 3/0 | no data | 32 channel | Axial/axial oblique | Yes | No |
| [**Terburg *et al.* 2012**](https://pubmed.ncbi.nlm.nih.gov/22832959/) | Hypervigilance for fear after basolateral amygdala damage in humans | GE-EPI | 42.9 | 3.5/0 | 4m 48s | no data | Axial/axial oblique* | Yes | Yes |
| [**Thorsen *et al.* 2019**](https://pubmed.ncbi.nlm.nih.gov/29753591/) | Emotion Regulation in Obsessive-Compulsive Disorder, Unaffected Siblings, and Unrelated Healthy Control Participants | GE-EPI | 39.3 | 2.8/0.2 | no data | no data | Axial/axial oblique | Yes | No |
| [**Tian *et al.* 2011**](https://pubmed.ncbi.nlm.nih.gov/22174900/) | Convergent evidence from multimodal imaging reveals amygdala abnormalities in schizophrenic patients and their first-degree relatives | GE-EPI | 47.3 | 4/0.8 | 7m | no data | Axial/axial oblique | Yes | No |
| [**Tong *et al.* 2019**](https://pubmed.ncbi.nlm.nih.gov/30653664/) | Real-time effects of interictal spikes on hippocampus and amygdala functional connectivity in unilateral temporal lobe epilepsy: An EEG-fMRI study | GE-EPI | 70.3 | 5/0 | 6m 46s | 8 channel | Axial/axial oblique | Yes | No |
| [**Torrisi *et al.* 2013**](https://pubmed.ncbi.nlm.nih.gov/23347587/) | Differences in resting corticolimbic functional connectivity in bipolar I euthymia | GE-EPI | 27 | 3/0.75 | 7m 2s | no data | Axial/axial oblique* | Yes | No |
| [**van Well *et al.* 2012**](https://pubmed.ncbi.nlm.nih.gov/22451349/) | Neural substrates of individual differences in human fear learning: evidence from concurrent fMRI, fear-potentiated startle, and US-expectancy data | GE-EPI | 15.8 | 3/0 | no data | 8 channel | Axial/axial oblique | Yes | No |
| [**Vasavada *et al.* 2021**](https://pubmed.ncbi.nlm.nih.gov/32900657/) | Effects of serial ketamine infusions on corticolimbic functional connectivity in major depression | GE-EPI | 8 | 2/0 | 6m 41s | 32 channel | Axial/axial oblique | Yes | No |
| [**Kumar *et al.* 2014**](https://pubmed.ncbi.nlm.nih.gov/25522395/) | Oxytocin affects the connectivity of the precuneus and the amygdala: A randomized, double-blinded, placebo-controlled neuroimaging trial | GE-EPI | 31.5 | 3.5/0 | 5m | 32 channel | Axial/axial oblique | Yes | No |
| [**Wackerhagen *et al.* 2017**](https://pubmed.ncbi.nlm.nih.gov/28294134/) | Influence of familial risk for depression on cortico-limbic connectivity during implicit emotional processing | GE-EPI | 27 | 3/1 | 4m 34s | no data | Axial/axial oblique* | Yes | No |
| [**Wackerhagen *et al.* 2020**](https://pubmed.ncbi.nlm.nih.gov/31637983/) | Amygdala functional connectivity in major depression - disentangling markers of pathology, risk and resilience | GE-EPI | 27 | 3/1 | 4m 34s | 12 channel | Axial/axial oblique* | Yes | No |
| [**Wang *et al.* 2009**](https://pubmed.ncbi.nlm.nih.gov/19427632/) | Functional and structural connectivity between the perigenual anterior cingulate and amygdala in bipolar disorder | GE-EPI | 42.2 | 3/0 | no data | no data | Axial/axial oblique | Yes | No |
| [**Wang *et al.* 2018**](https://pubmed.ncbi.nlm.nih.gov/29608759/) | Negative Schizotypy and Altered Functional Connectivity during Facial Emotion Processing | GE-EPI | 43.1 | 4/0 | 8m | no data | Coronal/coronal oblique | Yes | No |
| [**Wang *et al.* 2021**](https://pubmed.ncbi.nlm.nih.gov/32052123/) | The resting-state functional connectivity of amygdala subregions associated with post-traumatic stress symptom and sleep quality in trauma survivors | GE-EPI | 70.3 | 5/0 | 6m 48s | Standard | Axial/axial oblique* | Yes | Yes |
| [**Wei *et al.* 2021**](https://pubmed.ncbi.nlm.nih.gov/32710329/) | Effective connectivity predicts cognitive empathy in cocaine addiction: a spectral dynamic causal modeling study | GE-EPI | 36 | 4/0 | 6m | no data | Axial/axial oblique* | Yes | No |
| [**Westlund Schreiner *et al.*  2017**](https://www.ncbi.nlm.nih.gov/pmc/articles/PMC5555154/) | Multi-modal neuroimaging of adolescents with non-suicidal self-injury: Amygdala functional connectivity | GE-EPI | 8 | 2/0 | 5m 43s; 6m 28s | 32 channel | Axial/axial oblique | Yes | No |
| [**Wu *et al.* 2016**](https://www.ncbi.nlm.nih.gov/pmc/articles/PMC4837047/) | Age-related changes in amygdala-frontal connectivity during emotional face processing from childhood into young adulthood | GE-EPI; GE reverse spiral | 35.4; 70.3 | 3/0; 5/0 | 6m | 8 channel | Axial/axial oblique | Yes | No |
| [**Wu *et al.* 2019**](https://pubmed.ncbi.nlm.nih.gov/31357162/) | When worry may be good for you: Worry severity and limbic-prefrontal functional connectivity in late-life generalized anxiety disorder | GE-EPI | 12 | 3/0 | 3m 54s | 32 channel | Axial/axial oblique | Yes | No |
| [**Yang *et al.* 2021**](https://pubmed.ncbi.nlm.nih.gov/34335316/) | Abnormal Functional Connectivity of the Amygdala in Mild Cognitive Impairment Patients With Depression Symptoms Revealed by Resting-State fMRI | GE-EPI | 46.9 | 4.8/0 | no data | no data | Axial/axial oblique | Yes | No |
| [**Yin *et al.* 2011**](https://pubmed.ncbi.nlm.nih.gov/21813114/) | Altered resting-state functional connectivity of thalamus in earthquake-induced posttraumatic stress disorder: a functional magnetic resonance imaging study. | GE-EPI | 35.4 | 3/1 | no data | no data | Axial/axial oblique | Yes | No |
| [**Yuan *et al.* 2016**](https://pubmed.ncbi.nlm.nih.gov/27296506/) | Group cognitive behavioral therapy modulates the resting-state functional connectivity of amygdala-related network in patients with generalized social anxiety disorder | GE-EPI | 70.3 | 5/0 | 6m 48s | 12 channel | Axial/axial oblique | Yes | No |
| [**Zhang *et al.* 2017**](https://pubmed.ncbi.nlm.nih.gov/29066900/) | Abnormal functional connectivity of the posterior cingulate cortex is associated with depressive symptoms in patients with Alzheimer’s disease | GE-EPI | 46.9 | 4.8/0 | no data | 12 channel | Axial/axial oblique | Yes | No |
| [**Zhang *et al.* 2020**](https://pubmed.ncbi.nlm.nih.gov/32126520/) | Abnormal amygdala subregional-sensorimotor connectivity correlates with positive symptom in schizophrenia | GE-EPI^ | 39.1 | 4/0 | 7m | no data | Axial/axial oblique | Yes | Yes |
| [**Zhu *et al.* 2017**](https://pubmed.ncbi.nlm.nih.gov/26040546/) | Model-free functional connectivity and impulsivity correlates of alcohol dependence: a resting-state study | GE-EPI | 70.3 | 5/0 | no data | no data | Axial/axial oblique | Yes | No |
| [**Zich *et al.* 2020**](https://pubmed.ncbi.nlm.nih.gov/32574803/) | Modulatory effects of dynamic fMRI-based neurofeedback on emotion regulation networks in adolescent females | GE-EPI | 8 | 2/0 | 8m 54s; 4m 48s | 32 channel | Axial/axial oblique | Yes | No |
| [**Zielinski *et al.* 2018**](https://www.ncbi.nlm.nih.gov/pmc/articles/PMC6373177/) | Does development moderate the effect of early life assaultive violence on resting-state networks? An exploratory study | GE-EPI | 27; 22.5 | 3/0; 2.5/0.5 | 7m 30s | 8 channel;  32 channel | Axial/axial oblique | Yes | No |
| [**Zimmerman *et al.* 2019**](https://pubmed.ncbi.nlm.nih.gov/31396035/) | Functional brain changes during mindfulness-based cognitive therapy associated with tinnitus severity | GE-EPI | 18.8 | 3/0 | 10m; 3m 4s | 20 channel | Axial/axial oblique* | Yes | No |

^ Sequence type not stated but parameters in keeping with GE-EPI

* Imaging plane not stated but assumed to be axial/axial oblique
